# Decision support system to evaluate VENTilation in the Acute Respiratory Distress Syndrome (DeVENT study) – Trial Protocol

**DOI:** 10.1101/2021.08.25.21262610

**Authors:** Brijesh Patel, Sharon Mumby, Nicholas Johnson, Emanuela Falaschetti, Jorgen Hansen, Ian Adcock, Danny McAuley, Masao Takata, Dan S. Karbing, Matthieu Jabaudon, Peter Schellengowski, Stephen E. Rees, On behalf of the DeVENT study group

## Abstract

**Background:** The Acute Respiratory Distress Syndrome (ARDS) occurs in response to a variety of insults, and mechanical ventilation is life-saving in this setting, but ventilator induced lung injury can also contribute to the morbidity and mortality in the condition. The Beacon Caresystem is a model-based bedside decision support system using mathematical models tuned to the individual patient’s physiology to advise on appropriate ventilator settings. Personalised approaches using individual patient description may be particularly advantageous in complex patients, including those who are difficult to mechanically ventilate and wean, in particular ARDS.

**Methods:** We will conduct a multi-centre international randomised, controlled, allocation concealed, open, pragmatic clinical trial to compare mechanical ventilation in ARDS patients following application of the Beacon Caresystem to that of standard routine care to investigate whether use of the system results in a reduction in driving pressure across all severities and phases of ARDS.

**Discussion:** Despite 20 years of clinical trial data showing significant improvements in ARDS mortality through mitigation of ventilator induced lung injury, there remains a gap in its personalised application at the bedside. Importantly, the protective effects of higher positive end-expiratory pressure (PEEP) were noted only when there were associated decreases in driving pressure. Hence, the pressures set on the ventilator should be determined by the diseased lungs’ pressure-volume relationship which is often unknown or difficult to determine. Knowledge of extent of recruitable lung could improve the ventilator driving pressure. Hence, personalised management demands the application of mechanical ventilation according to the physiological state of the diseased lung at that time. Hence, there is significant rationale for the development of point-of-care clinical decision support systems which help personalise ventilatory strategy according to the current physiology. Furthermore, the potential for the application of the Beacon Caresystem to facilitate local and remote management of large numbers of ventilated patients (as seen during this COVID-19 pandemic), could change the outcome of mechanically ventilated patients during the course of this and future pandemics.

**Trial registration:** ClinicalTrials.gov identifier (*NCT* number): NCT04115709

**Administrative information:** Note: the numbers in curly brackets in this protocol refer to SPIRIT checklist item numbers. The order of the items has been modified to group similar items (see http://www.equator-network.org/reporting-guidelines/spirit-2013-statement-defining-standard-protocol-items-for-clinical-trials/).

|  |  |
| --- | --- |
| Title | **Decision support system to evaluate **VENT**ilation in the Acute Respiratory Distress Syndrome (DeVENT study)** |
| Trial registration | ClinicalTrials.gov Identifier: NCT04115709 |
| Protocol version | 11<sup>th</sup> May 2021, version 4.0 |
| Funding | This study is fully funded by the European Commission |
| Author details | Dr Brijesh Patel<br>Dr Sharon Mumby<br>Mr Nicholas Johnson<br>Dr Emanuela Falaschetti<br>Mr Jorgen Hansen<br>Professor Ian Adcock<br>Professor Danny McAuley<br>Professor Masao Takata<br>Dr Dan S. Karbing<br>Professor Matthieu Jabaudon<br>Dr Peter Schellengowski<br>Professor Stephen E. Rees |
| Name and contact information for the trial sponsor | Imperial College London<br>Joint Research Compliance Office<br>Imperial College London and Imperial College Healthcare NHS Trust<br>Room 215, Level 2, Medical School Building<br>Norfolk Place<br>London, W2 1PG<br>/ 0207 594 1862 |
| Role of sponsor | The study sponsor has overseen the design of the study and will have oversight of the trial. The sponsor has ensured that the trial protocol, Patient Information Sheet (PIS), Informed Consent Form (ICF), letter and submitted supporting documents have been approved by a Research Ethics Committee (REC) within each country, prior to any patient recruitment taking place. This study will be conducted in compliance with the protocol approved by the REC and according to GCP standards, UK and local Clinical Trials Regulation.<br>Data ownership rights will lie with the institution. Our expectation is that after data analysis, information from this study will be widely disseminated in the medical and scientific community. |

## Introduction

### Background and rationale

Despite 20 years of clinical trial data showing significant improvements in ARDS mortality through lung protective ventilation via low tidal volume ventilation (1), limitation of inspiratory pressures (2), conservative fluid management (3), open lung ventilation (4), and prone position (5), there remains a gap in the personalised application at the bedside. Needham et al showed 69% of ventilator settings are non-adherent to lung protective ventilation strategies (6). A recent study has shown that non-personalised recruitment of the lung utilising high pressures leads to harm (7). A machine learning analysis of the data from this study showed that this negative impact was greater in a proportion of patients with consolidated lung with pneumonia (likely to be non-recruitable) (8). In addition, a ventilator driving pressure of >16cmH_2_O is associated with a significant increase in mortality (2). In this study, Amato and colleagues showed a strong association between driving pressure and survival even though all the ventilator settings that were used were lung-protective. Importantly, the protective effects of higher PEEP were noted only when there were associated decreases in ΔP. Hence, the pressures set on the ventilator should be determined by the diseased lungs’ pressure-volume relationship which is often unknown or difficult to determine. Knowledge of the extent of recruitable lung could improve the ventilator driving pressure. Hence, personalised management demands the application of mechanical ventilation according to the physiological state of the diseased lung at that time. Hence, there is significant rationale for the development of point-of-care clinical decision support systems which help personalise ventilatory strategy according to the current physiology.

The Beacon Caresystem (9,10) is a model-based bedside decision support system using mathematical models tuned to the individual patient’s physiology to advise on appropriate ventilator settings. Personalised approaches using individual patient description may be particularly advantageous in complex patients, including those who are difficult to mechanically ventilate and wean, in particular ARDS. The core of the system is a set of physiological models including pulmonary gas exchange, acid-base chemistry, lung mechanics, and respiratory drive (10).

The Beacon Caresystem tunes these models to the individual patient such that they describe accurately current measurements of lung physiology to base further clinical decisions and monitor lung health in critical care. Once tuned, the models are used by the system to simulate the effects of changing ventilator settings. The results of these simulations are then used to calculate the clinical benefit of changing ventilator settings by balancing the competing goals of mechanical ventilation. For example, an increased inspiratory volume will reduce acidosis of the blood while detrimentally increasing lung pressure. Appropriate ventilator settings therefore imply a balance between the clinical preferred value of pH weighted against the preferred value of lung pressure. A number of these balances exist, and the system weighs these, calculating a total score for the patient for any possible ventilation strategy. The system then calculates advice as to changes in ventilator settings to improve this score.

The Beacon Caresystem includes a number of physiological models, each of which have been validated separately, and as part for the Beacon Caresystem. Gas exchange parameters are estimated from a procedure varying inspired oxygen fraction, known as the Automatic Lung Parameter Estimation (ALPE) procedure. Mathematical models of pulmonary gas exchange have been shown to describe patients during and following surgery (11)(12)(13) in the ICU (14-16), and have been validated against the experimental reference technique for measuring gas exchange (17)(18). Modelling of acid-base levels have been shown to accurately simulate changes in CO_2_, O_2_ and strong acid in blood (19), as well as the mixing of blood from different sources (20) whilst models of respiratory drive accurately simulate the effects of changes in support ventilation (21). The Beacon Caresystem has been shown in 4-8 hour periods to reduce levels of inspired oxygen, tidal volume, and pressure support, without detrimental effects (22) and while protecting the respiratory muscles (23).

The advice provided by the system is presented to the clinician. The ventilator settings are then changed by the clinician, and the patient’s physiological response to these changes is automatically used by the system to re-tune the models and repeat the process of generating new advice. Patients with severe lung abnormalities such as acute respiratory distress syndrome (ARDS), which often result in small, stiff lungs, are often in control ventilation mode with little or no spontaneous breathing. For these patients, the clinician often increases PEEP to try to recruit units of the lung which are collapsed. This can be difficult, as increasing PEEP may result in elevated lung pressure and hence an increased risk of lung injury, incomplete expiration and air trapping, and haemodynamic compromise, especially in those with heart failure. The Beacon Caresystem will advise the clinician to optimise PEEP according to the measured physiology at that time and hence, will enable the clinician to determine if the patient is a responder or non-responder to the PEEP manoeuvre. PEEP optimisation should enable best ventilation according to personalised press-volume assessments with potential reductions in driving pressure. Importantly, risk is mitigated by the fact that the clinician can always override the advice given.

#### Case for understanding physiological trajectory and response

In parallel, the prospective, longitudinal analysis of physiological trajectory, and physiological responsiveness to adjunctive manoeuvres (e.g. prone position, recruitment manoeuvre, fluid balance) during ARDS is also of key importance. Such a study if embedded in a randomised clinical trial would enable robust data to discover the timely prediction of deterioration and to discern the optimal time for the application of more invasive interventions such as ECMO. At present there is no clinically validated lung physiology monitor to longitudinally assess breath by breath respiratory physiology in ARDS.

#### COVID-19 specific aspects

Critical cares across the world have admitted numerous mechanically ventilated patients with COVID-19 pneumonia. ARDS secondary to COVID-19 has been thought to present with a different respiratory physiology and trajectory as compared to non-COVID19 induced ARDS. We will include a sub-analysis to test the Beacon Caresystem in the ventilatory management of patients specifically admitted with COVID-19-induced ARDS but also allow a deeper physiological understanding of COVID19-induced ARDS. In addition, this will include a comparison with those not effected by COVID-19 or mixed, for instance, respiratory compliance, dead space, shunt fraction. Both would enable mapping of trajectory and application of personalised therapies through physiological enrichment. Finally, a beneficial effect from the Beacon Caresystem with its immediate application to facilitate the local and remote management of large numbers of ventilated patients (as seen during this pandemic), could change the outcome of mechanically ventilated patients during the course of this and future pandemics.

### Objectives

**RCT Primary objectives and outcome (in all sites): The overall final analysis will be performed as the average driving pressure per unit time. However, given the nature of ICU management (transfer, disconnection, nebulisation etc), the driving pressure calculation will only involve periods when the Beacon Caresystem is operating and attached successfully to the patient**.

### RCT Secondary objectives/outcomes (in all sites)

1. Daily average calculated delivered pressure over time, for periods of spontaneous breathing.
2. Daily average calculated mechanical power over time.
3. Daily average calculated oxygenation index over time.
4. Daily average ventilatory ratio over time.
5. Ventilator free days at 90 days.
6. Time from control mode to support mode.
7. Number of changes in ventilator settings per day.
8. % of time in control mode ventilation.
9. % of time in support mode ventilation.
10. Total duration of mechanical ventilation.
11. Tidal volume over time.
12. PEEP setting over time.
13. Ventilation related complications e.g. pneumothorax and/or pneumomediastinum
14. Device malfunction event rate
15. Device related adverse event rate
16. Number of times the advice from the Beacon system is followed through the duration of the study.

### Trial design

The DeVENT study is a 2.5 year multi-centre international randomised, controlled, allocation concealed, open, pragmatic clinical trial which will enrol patients with ARDS (according to Berlin definition). Patients will be randomised to either have the Beacon Caresystem attached with advice activated (N=55) or standard care with the Beacon Caresystem attached with advice inactive (N=55). The study will also use the Beacon Caresystem alongside sequential biological sampling to deep phenotype patients with ARDS.

## Methods: Participants, interventions and outcomes

### Study setting

DeVENT is recruiting subjects with ARDS prospectively from four clinical ICU sites across Europe, namely the Royal Brompton Hospital, UK; Harefield Hospital, UK; Medical University Vienna (MUV), Austria; and Clermond-Ferrand University Hospital, France.

### Eligibility criteria

The inclusion criteria are:

- Invasive mechanical ventilation.
- A known clinical insult with new worsening respiratory symptoms
- Chest radiograph with bilateral infiltrates consistent with evidence of pulmonary oedema but not fully explained by cardiac failure.
- Hypoxaemia as defined by PaO2/FiO2 of ≤300mmHg (or ≤40kPa) (pre-ECMO PaO_2_/FiO_2_ will be used should patient be placed on extracorporeal support).

Patients will be excluded if any of the following are met:

- Age < 18 years old.
- The absence of an arterial catheter for blood sampling at study start.
- Consent declined.
- Over 7 days of mechanical ventilation.
- Treatment withdrawal imminent within 24 hours.
- DNAR (Do Not Attempt Resuscitation) order in place
- Severe chronic respiratory disease requiring domiciliary ventilation and/or home oxygen therapy (except for sleep disordered breathing)
- Veno-Arterial ECMO
- Head trauma or other conditions where intra-cranial pressure may be elevated and tight regulation of arterial CO2 level is paramount

### Who will take informed consent?

Consent to enter the study must be sought from each participant only after a full explanation has been given, an information leaflet offered and time allowed for consideration. Signed participant consent should be obtained. The right of the participant to refuse to participate without giving reasons must be respected. In most cases it will not be possible to gain prospective consent for the patient at the time of enrollment. The nature of the study’s population is such that patients are critically ill and often unconscious and, in many cases, will not be able to grant consent themselves. As the Beacon Caresystem device takes an open loop approach with the decision to change the ventilator settings ultimately lying with the clinical team, there is minimal risk in participation into the study. Hence, given the emergent nature of the study situation the Beacon Caresystem (with the advice turned off) will be attached to the patient and samples collected. Given the urgent nature of COVID-19 and the process of ECMO, at the discretion of the clinical team, prior to consent/advice below a similar sample of blood may be taken prior to starting ECMO (T0) and hence, often prior to discussion with a personal nominee. If they are not enrolled into the study then this sample will be discarded. No analysis of data or samples will occur until consent/advice as below is obtained. This has been discussed with our Patient and Public Involvement (PPI) representative who is fully supportive of this process.

#### Personal legal representative advice

Due to the acute care trial setting and the vulnerability of the patients’ population, a patient information sheet will be provided and advice sought from a third party acting as a consultee; in most cases this person will be a personal consultee, who is someone who knows the person lacking capacity and is able to advise the researcher about that person’s wishes and feelings in relation to the project and whether they should participate in the research. This person must be interested in the welfare of the patient in a personal capacity, not in a professional capacity or for remuneration and will most likely be a relative, partner, legal power of attorney, or close friend. Written advice will be documented via the personal consultee declaration form and stored in both the medical notes and the site file. However, where the personal consultee is not available on site, the researcher may contact the personal consultee via telephone and seek verbal advice. The researcher will talk the consultee through the patient information sheet and send a copy via email or in the mail. The verbal agreement will be recorded in the telephone consultee declaration form. The telephone consultee declaration form will be signed by a second member of staff who has witnessed the telephone advice. This witness may be a member of the research team or site medical staff. A copy of the telephone consultee declaration form will be placed in the medical notes and the site file. In such cases, where a telephone consultation occurs, a written personal consultee declaration form will be obtained as soon as possible.

#### Professional legal representative advice

Where no Personal Consultee is available, the researcher will nominate a professional person (independent of the study) to assist in determining the participation of a person who lacks capacity. The nominated professional consultee is someone who will be appointed by the researcher to advise the researcher about the person’s (who lacks capacity) wishes and feelings in relation to the project and whether they should participate in the research. In the event a Personal Consultee cannot be identified the appointed Nominated Consultee will not be involved in either the research project or the patient’s clinical care and will be completely independent of both. In the event that a personal consultee is identified after the nominated professional consultee advice has been obtained, the above process for personal consultee declaration will be followed and all advice forms will be filled as detailed previously.

#### Retrospective patient consent

If a surviving patient regains competency, we will approach the patient to obtain their consent to continue in the study. This will be obtained via telephone, in the event that the patient cannot attend follow up visits. If the patient refuses consent or prefers to withdraw during this ICU stay the intervention will be stopped but the regular/expected medical care will still be provided. We will ask the patient if we can use their existing hospital data. Without their consent, no additional information about the patient will be collected for the purposes of the study. However, to maintain integrity of the randomized trial, all information collected up to that time will still be used and analyzed as part of the study.

A copy of the signed Informed Consent Form (ICF) along with a copy of the most recent approved Patient Information Sheet (PIS) will be given to the study participant. The original signed consent form will be retained at the study site (one filed in the medical notes and one filed in the Investigator Site Master File (ISF)). A copy of the consent form will also be given to the patient. If new safety information results in significant changes to the risk–benefit assessment, the consent form will be reviewed and updated if necessary. All subjects, including those already being treated, will be informed of the new information, given a copy of the revised consent form and asked to re-consent if they choose to continue in the study.

### Additional consent provisions for collection and use of participant data and biological specimens

Biological samples including blood, urine, broncho-alveolar lavage/brushings will be stored in a biobank for future analysis. All analyses undertaken will relate to furthering the understanding of ARDS. These samples will be identified only by a numerical identifier and results from these tests including genetic information will not be stored in the case notes or given to the subject, their family or doctors involved with their care. The samples will be stored in an approved secure facility. Only approved researchers will be able to access the samples. The sample labels will contain no subject identifiable information. The movement and storage of samples will be carried out in accordance with the Human Tissue Act and respective national regulations. Further details are provided in Appendix 3.

### Interventions

#### Explanation for the choice of comparators

In the comparator group, mechanical ventilation is managed according to standard care at the local centre. In this group Beacon advice will be disabled, with the system being used solely for data collection including changes in ventilation. Physiological variables captured in this arm of the study will mirror the intervention arm. The Beacon will remain on the patient for as long as the patient stays in the study centre, is successfully extubated (i.e. not requiring invasive ventilation or nasal high flow oxygenation for >24 hours), attains trachemask for >24 hours, or passes away. In addition, ALPE measurements will be perfomed as per intervention arm. This strategy will allow for comparison of physiological status between control and intervention arms, but will not risk modifying care in the control arm.

#### Intervention description

Once enrolled into the study and randomized to intervention, the Beacon will provide advice including changes in ventilation and measurement of parameters including shunt fraction and ventilation/perfusion mismatch (using an automated lung parameter estimation or ALPE procedure) to the treating clinician. Should the patient go onto a mode not supported by the Beacon Caresystem (e.g. Airway Pressure Release Ventilation (APRV)) or onto ECMO, the Beacon Caresystem advice will be paused. Once a supported mode is re-established the Beacon advice will be reactivated. The Beacon will remain on the patient for as long as the patient stays in the study centre, is successfully extubated (i.e. not requiring invasive ventilation or nasal high flow oxygenation for >24 hours), attains trachemask for >24 hours, or passes away.

In addition to when the Beacon provides ventilatory advice, standard clinical procedures will be performed either to place the patient in prone position, or perform recruitment manoeuvres (figure 2). For each of these procedures, the ALPE procedure will be performed prior to, during and subsequent to the start and end of the procedure (figure 2). However, due to the nature of ICU processes in some centres looking after COVID-19 patients, this may not be possible.

**Figure 1:**
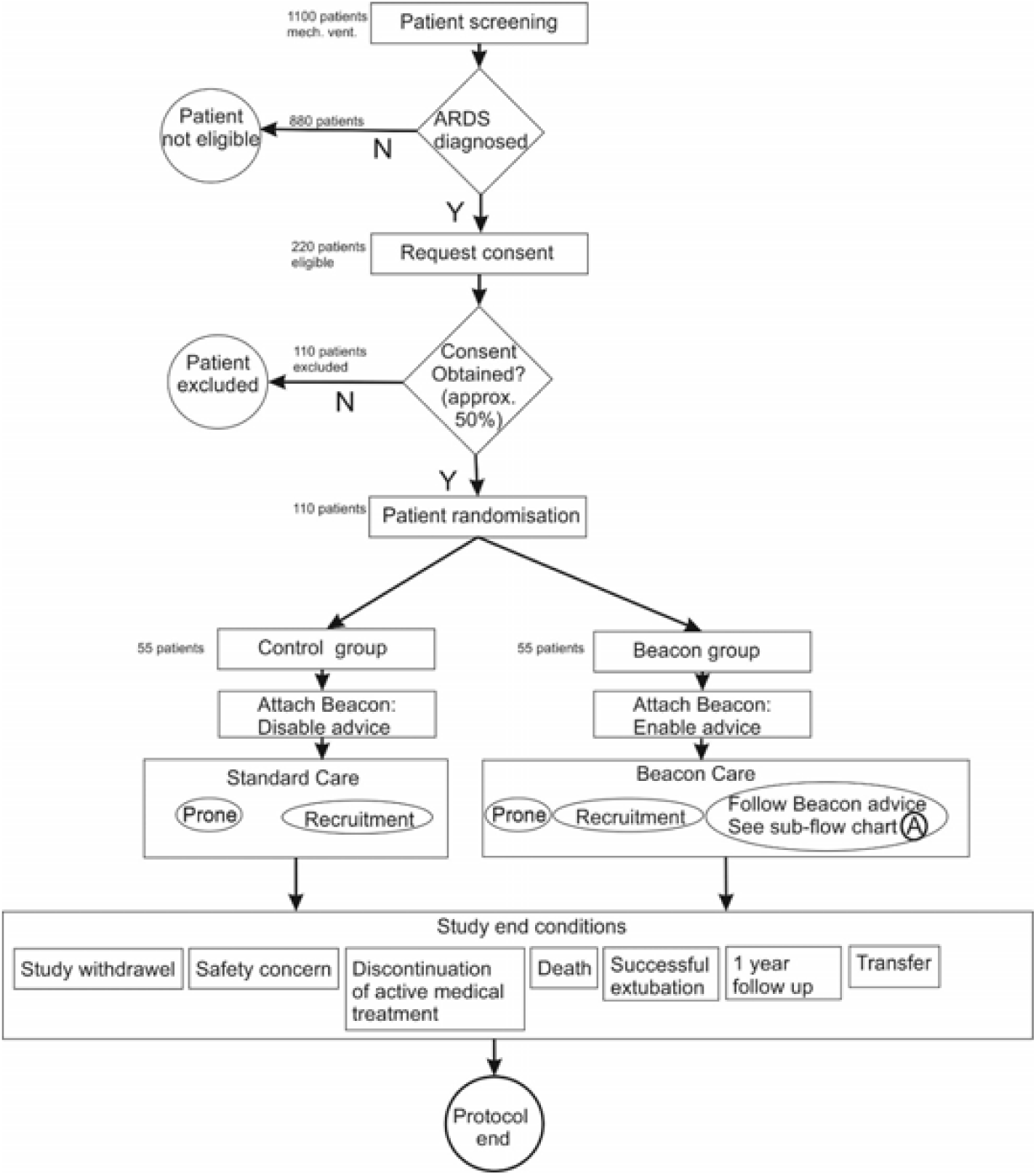
A randomised, controlled, allocation concealed, open, pragmatic clinical study investigating the effect of the Beacon Caresystem on ventilation in ARDS.

**Figure 2.**
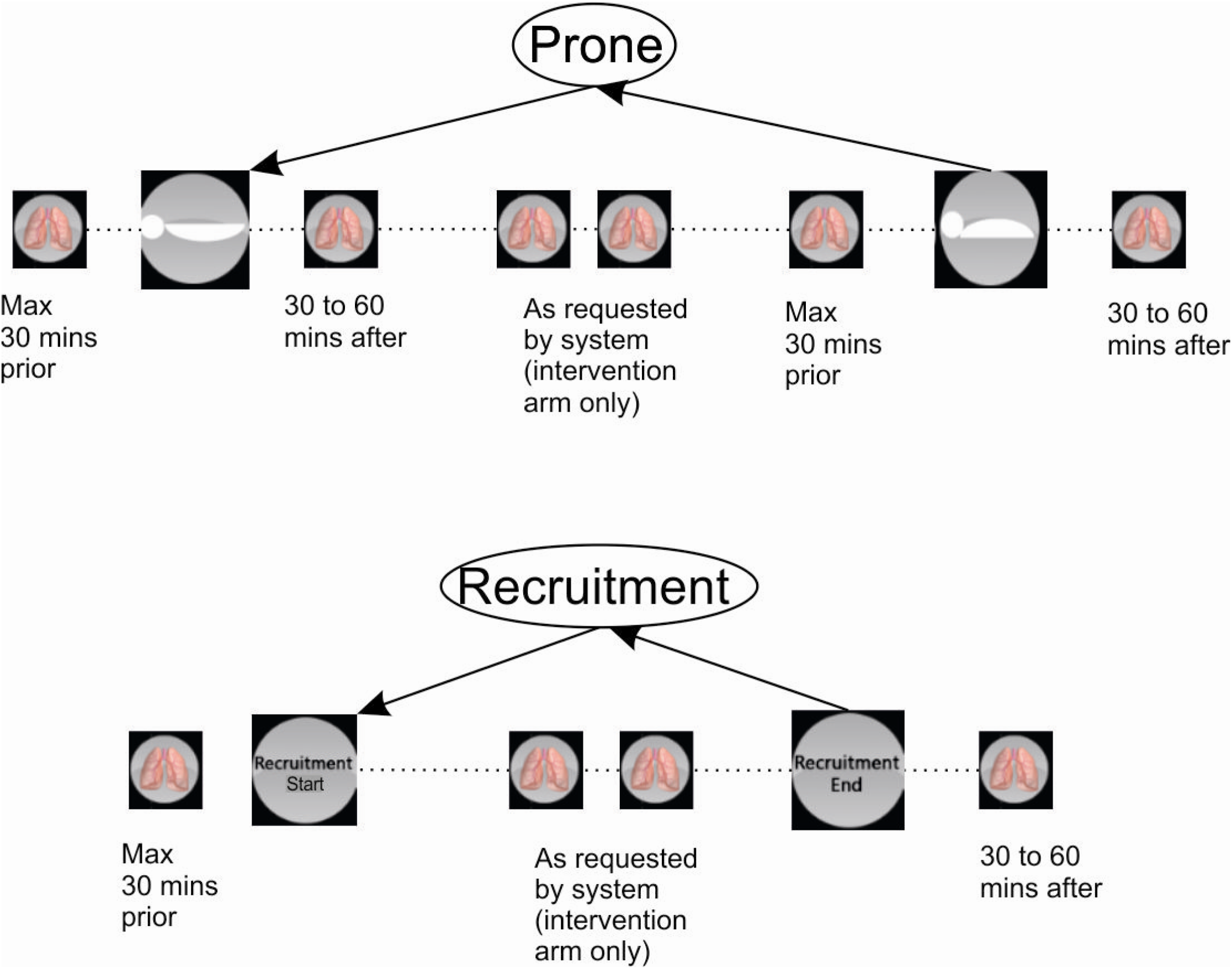
Prone position and recruitment manoeuvre flow, including intermittent ALPE procedures.

### Criteria for discontinuing or modifying allocated interventions

Patients randomised on ECMO or placed on ECMO post-randomisation will be attached to the Beacon Caresystem but the system will only be activated to give advice during periods when the ECMO sweep gas is off. During such periods, the interventions described above as per control and intervention arms will be followed. For instance, once or twice daily routine ALPE procedures will be performed with at least 8 hours duration between these routine ALPE measurements. In addition, an ALPE procedure will also be performed as soon as possible after sweep has been switched off and advice will be activated in the intervention arm. However, the ALPE will not lead to advice or be shown in both control or intervention arms as advice will not be on when ECMO gas sweep is on. If ventilation mode that is not supported by the Beacon Caresystem is commenced, the advice of the system will be paused and restarted once a compatable mode has been reestablished.

### Strategies to improve adherence to interventions

The researcher will check connectivity of the device over the course of the period. To assess compliance, the number of ventilatory decisions made over time will be compared to the number advised by the Beacon Caresystem.

### Relevant concomitant care permitted or prohibited during the trial

None applicable

### Provisions for post-trial care

None applicable

### Outcomes

#### RCT Primary objectives and outcome (in all sites)

##### Objective

To assess the average driving pressure delivered by the mechanical ventilator over the period of time when ARDS ventilation management is advised by the Beacon Caresystem as compared to standard care.

*Driving pressure will be measured once a day, using end inspiratory and expiratory pauses. Respiratory pressures at the end of inspiratory (Pplat) and expiratory (PEEP) pauses are known to approximate average pressure in the alveoli at these points, such that their difference, Pplat-PEEP, is the correct measure of driving pressure applied to the lungs. This measurement will only be performed in breaths where no spontaneous breathing activity occurs. In addition to the measurement of driving pressure, surrogate measurements of driving pressure will be obtained continuously by approximating Pplat with peak inspiratory pressure (Ppeak), and PEEP with PEEP values set on the ventilator (PEEPset)*.

##### Outcome

The overall final analysis will be performed as the average driving pressure per unit time. However, given the nature of ICU management (transfer, disconnection, nebulisation etc), the driving pressure calculation will only involve periods when the Beacon Caresystem is operating and attached successfully to the patient.

#### RCT Secondary objectives/outcomes (in all sites)

##### Objective

In mechanically ventilated patients with ARDS we will determine the effects of the Beacon Caresystem compared to standard care on the outcomes listed below.

1. *Daily average calculated delivered pressure over time, for periods of spontaneous breathing.
2. *Daily average calculated mechanical power over time.
3. *Daily average calculated oxygenation index over time.
4. *Daily average ventilatory ratio over time (33).
5. Ventilator free days at 90 days.
6. *Time from control mode to support mode.
7. *Number of changes in ventilator settings per day.
8. *% of time in control mode ventilation.
9. *% of time in support mode ventilation.
10. *Total duration of mechanical ventilation.
11. *Changes in tidal volume over time.
12. *Changes in PEEP setting over time.
13. Ventilation related complications e.g. pneumothorax and/or pneumomediastinum
14. Device malfunction event rate
15. Device related adverse event rate
16. Number of times the advice from the Beacon system is followed through the duration of the study.

*\*Data used for these analyses will only be taken from non-ECMO times*.

### Exploratory objectives/outcomes

See supplementary appendix

### Participant timeline

Patients will be admitted in ICU and will have a routine clinical follow up visit.

### Sample size

The original required sample size for the primary outcome is 110 patients. 55 patients per group will allow detection of a difference of 2 cmH_2_O in driving pressure between the groups with 90% power and a two-sided alpha of 0.05 assuming a control group driving pressure of 15 cmH2O with a standard deviation of 2.5 cmH2O and including a 40% dropout (33 patients per group for analysis). We have used data from the MIMIC dataset (as published in Serpa Neto et al (25)) for the estimation of the driving pressure. In view of the longitudinal analysis, loss to follow up has taken account of an average mortality of 34% and a 6% drop out.

A recalculation of sample size was undertaken in January 2021 (in view of the COVID-19 pandemic) to evaluate whether the extent of driving pressure data being collected was sufficient to answer the primary objective. This was due to a higher proportion of patients being exclusively on ECMO than expected due to the COVID-19 as well as potential differences in physiology between COVID-19 and non-COVID-19 ARDS.

Basing the calculation on a repeated-measures basis using estimates of the intraclass correlation coefficient (ICC) of driving pressure values and the estimate of the coefficient of variation (CV) for days data collected (per patient) we calculated the expected patients required to fulfil the primary outcome. Using the data extract collected for the second Data Monitoring and Ethics Committee (DMEC) meeting (December 2020, n=38) we estimated values for ICC and CV of 0.3 and 0.8 respectively. Based on these estimates and by providing alternative scenarios we were able to create the following table:

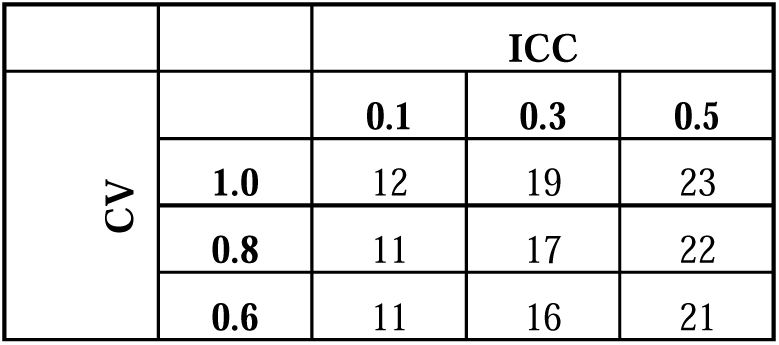

|  |  | ICC |  |  |
| --- | --- | --- | --- | --- |
|  |  | 0.1 | 0.3 | 0.5 |
| CV | 1.0 | 12 | 19 | 23 |
|  | 0.8 | 11 | 17 | 22 |
|  | 0.6 | 11 | 16 | 21 |
|  | 0.4 | 11 | 15 | 20 |

From the above, under the ‘worst-case’ scenario; with a CV of 1.0 and an ICC of 0.5 we would require 23 patients per arm to provide sufficient driving pressure data to have a powered analysis. Based on the December 2020 data it is estimated that approximately 96 patients would be required for a powered analysis. A further investigation (February 2021, n=65) confirmed that the estimates for ICC and CV were appropriate and were unlikely to go beyond the extreme values presented above.

Comparing this to the original calculation and taking the increased rate of ECMO patients into consideration, it is expected that the original proposal to randomise 55 patients per group would allow us to collect sufficient driving pressure data to answer the primary objective.

### Recruitment

The required sample size for the primary outcome is 96 patients across four ICU sites.

### Assignment of interventions: allocation

#### Sequence generation

Patients will be randomly allocated to one of two groups, i.e. the Standard Care group or the Beacon group. Randomisation will be stratified by site; ECMO/non ECMO; and COVID/non-COVID. Patients will be centrally allocated to an arm of the study from a master randomisation list created by the study statistician, stratified by ECMO and COVID-19 status to control for different patient severity across parts of the year. Given the unblinded nature of this device study, allocation concealment will be maintained through random size block allocation of patients.

### Concealment mechanism

See 16a

### Implementation

See 16a

### Assignment of interventions: Blinding

**Who will be blinded**

This is an unblinded study.

### Procedure for unblinding if needed

N/A

### Data collection and management

#### Plans for assessment and collection of outcomes

DeVENT study data are entered directly into the SMART-TRIAL database and respiratory physiological data will be collected through the beacon Caresystem. Figure 3 shows the Data workflow process. The principle means for data collection will be through electronic data capture through the internet on the SMART Trial database by each individual study site. Each participant will be allocated a unique Participant Study Number at trial entry, and this will be used to identify the individual on the CRF for the duration of the trial. Data will be collected from the time of trial entry until hospital discharge. Trial data will be entered onto a CRF and processed electronically as per ICTU Standard Operating Procedures (SOPs) and the study specific Data Management Plan. Data queries will be raised electronically. Data will be collected in a pseudo anonymous form on the Beacon Caresystem (measurements by the mechanical ventilator and measurements by Beacon sensors of respiration, gas exchange, lung mechanics, metabolism and entered arterial blood gas results). This breath by breath raw data collected on the Beacon Caresystem will be securely transferred, stored and processed at Aalborg university. Both eCRF and the Beacon Caresystem will contain only coded reference to each subject. Data will be collected from the device after each patient by the research team and with the assistance from clinical engineering support to ensure each patient’s data is fully saved on a secure cloud database. The volume of data (breath by breath recording) means that it cannot be recorded on any CRF and University of Aalborg will generate an automatic report per patient. Data will be entered in the CRF in cases of adverse events related and not related to mechanical ventilation.

**Figure 3.**
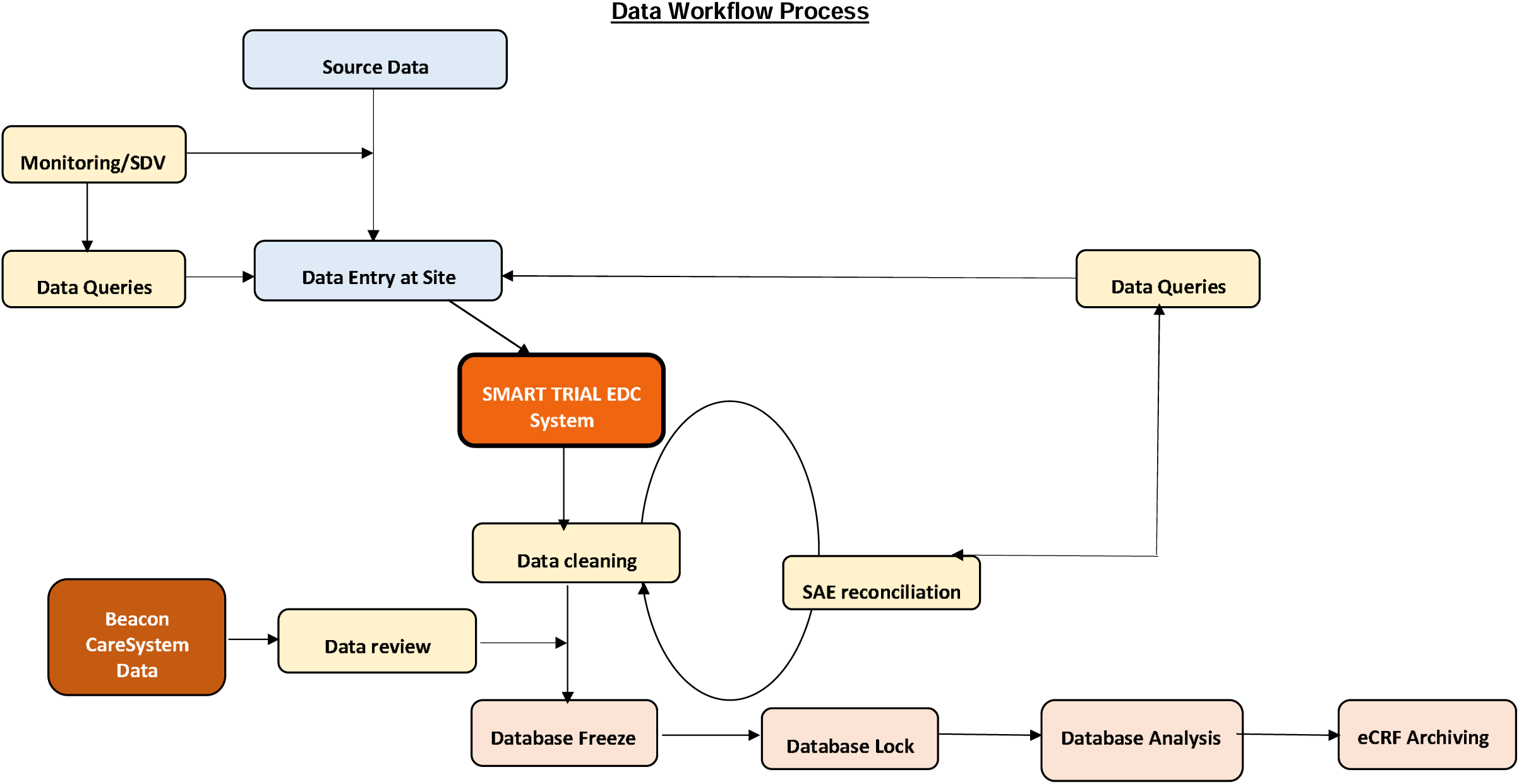
Data workflow process.

All additional research patient data will be completed in an electronic Case Report Form (eCRF) which will be configured within the ICCA database (electronic patient record) used in each of the intensive care units under standard operating procedures.

Data will be managed according to the NHS Act 2006, the Health and Social Care Act 2012, the Data Protection Act 2018, and the Human Rights Act 1998 and the study will be conducted according to UK code of Good Clinical practice (GCP) for research.

### Plans to promote participant retention and complete follow-up

In light of the COVID-19 pandemic the protocol was modified to minimize the number of face to face visits. A phonecall discussion with personal nominees was initiated and retrospective phone consent for patients that survive was also included. Postal questionaires were also implemented.

### Data management

Data entry of the eCRF is the responsibility of the study site. Data are provided manually (data entry). Data entry is performed by site-specific users. System queries will automatically be generated by the SMART-TRIAL system. Data management activities are conducted via the eCRF. Manually entered data are validated, queried (if necessary) and cleaned. Once the eCRFs have been reviewed by the Study Manager/Monitor, and any queries raised, the Study Manager/Monitor or designee will perform data cleaning and data review issuing additional queries on any discrepant data or where further clarification is required in relation to system generated queries. The site will answer the queries updating the eCRF data when appropriate. The Study Manager/Monitor or designee will close system and manual queries. Monitoring and Source Data Verification (SDV) will be performed electronically via the eCRF by the Study Manager/Monitor as defined in the study monitoring plan.

External data (data collected outside of the eCRF) will be provided electronically. The details regarding the sources and types of external data collected for this study, as well as reconciliation and integration processes are specified in Section 19. Once the eCRF forms/screens are declared clean (no outstanding queries or issues) and locked by the Study Manager/Monitor at the end of the study the CI must complete the signature panel. The Study Manager will request CI approval and then proceed with Database Lock (DBL). Data entry into the eCRF is the responsibility of the study site staff. The study site team will create database records for every enrolled subject via the SMART-TRIAL system by entering the eCRF data directly into the SMART-TRIAL database.

Prior to subject being enrolled on the database, the subject ID and Randomisation number/code for each subject will be allocated manually by the Study Manager. The Data Manager will perform a quality control check of the Master randomisation list to ensure accuracy and completeness of entries. Prior to database lock, the Study Manager/Monitor will confirm that all subject data has been entered and source data verified in accordance with the study Monitoring Plan.

### Confidentiality

The investigator must ensure that the subject’s confidentiality is maintained. On the CRF or other documents submitted to the Sponsors, subjects will be identified by a subject ID number only. Documents that are not submitted to the Sponsor (e.g. signed informed consent form) should be kept in a strictly confidential file by the investigator. The investigator shall permit direct access to subjects’ records and source documents for the purposes of monitoring, auditing, or inspection by the Sponsor, authorised representatives of the Sponsor, NHS, Regulatory Authorities and RECs.

### Plans for collection, laboratory evaluation and storage of biological specimens for genetic or molecular analysis in this trial/future use

#### Bio- and image-banking procedures

The scope is to cover all human samples and associated data in the study. The Patient Informed Consent also covers many of the details described here. The types of samples acquired in this study are described above, and includes samples of peripheral blood, urine, bronchial lavage, and bronchial brushes. These samples will be processed from fresh into different formats, for storage at different temperatures. All sample management (acquisition, processing, storage, shipment, analysis, disposal) throughout the study will be in accordance with:

- EU and relevant national legal and ethical standards, (including, any national research governance requirements and self regulation). The investigators in the project will provide the Management and Ethics Board with a summary of their applicable national or institutional rules regarding Biobank Activities, as well as changes national legal requirements related to Biobank Activities.
- the procedures detailed in the Study Reference Manual (SRM)

The SRM will be provided to all participating centres. All samples will be managed only by fully trained and qualified research personnel. Images will be stored in the Royal Brompton & Harefield NHS Foundation Trust servers as these images may contain useful information for clinical management of the patient. Images will be performed or analysed in a blinded manner to ensure allocation concealment. If images are transferred outside hospital servers, they will be pseudoanonymised using the study number.

#### Coding of samples and data

Special precautions will be taken to ensure the study is carried out with a high degree of confidentially. All study data, images, and samples related to the subject will be coded. Each subject will be allocated a code number at the time consent is given. The code will be used to identify him/her and associated data and results, without having to use his/her name, medical record number, or other common identifiers. The coding of all information resulting from the subject’s participation in the study is to ensure that the results are kept confidential by keeping the subject’s identity and results separate. Each sample will have a sample identifier. Only the coded information will leave the investigator site. No information will be stored on the sample labels that may be used to identify the subject. The subject code together with the sample identifier ensures each sample is uniquely identified. Only the clinical investigator at each site will have access to the code key with which it is possible to connect personal data to the individual participant. These data are available for review by the sponsor monitoring the study, regulatory authorities and independent ethics committees. The purpose of these reviews is to assure the proper conduct of the study. For the blood sample for genetic analysis extra precautions will be taken to ensure confidentiality. The sample will be labelled with the same code that is given to the subject in the study. As an added level of security, the DNA when it is extracted from the blood sample and the results of any research on the DNA will also receive a second code number. A file linking the first and second codes will be kept in a secure place with restricted access. If the subject changes his/her mind about participating in the genetic research, this link will allow the sample to be located and destroyed.

#### Sample and image data storage

Samples taken for each subject will either be sent for immediate analysis, local to the investigator site, to a specialised analysis centre, or processed further and stored at the investigator site, in the local storage facilities before dispatch to, and storage in facilities at Imperial College London.

All samples and image data will be stored securely, with access restricted to approved staff, until required for analysis. The Designated Individual at Imperial College London will be accountable to the Human Tissue Authority for compliance with the Human Tissue Act, the local Person Designated will be responsible for the samples.

All storage facilities will have their own local contingency and disaster/recovery plans. The investigator, for each investigator site, will be the local custodian. He/she will be, accountable for the safe-keeping of the samples and associated data, unless otherwise specified by law. The location of each sample and image will be recorded and tracked throughout its life cycle to maintain a chain-of-custody, in accordance with the SRM. At the end of the storage time any remaining samples and images will be destroyed and their disposal recorded, unless otherwise required to be anonymised by local ethics. As new scientific data become available we will be able to use this resource of stored samples and images to investigate if this new data is relevant pending additional ethical approval. Samples and images will be stored for up to 20 years. No staff at any storage facilities or facilities outside clinical work areas will have information that directly identifies any subject.

### Statistical methods

#### Statistical methods for primary and secondary outcomes

Detailed analysis methods is documented in the study’s Statistical Analysis Plan (SAP) (see supplementary appendix). Any deviations from the statistical analysis plan will be dealt with in accordance with Imperial Clinical Trial Unit’s statistical Standard Operating Procedures.

In summary, the primary outcome of driving pressure during ventilation and the other repeated measurements outcomes will be analysed using a mixed model, including covariates for site and stratification variables, duration of ventilation prior to Beacon connection, duration of hospital admission prior to intubation and number of non-ECMO, non-support-mode days. Categorical data will be presented as number and percentage and any comparisons between the two groups completed using the chi-squared or Fishers exact test. Logistic regression will be used where adjustments for important covariates are required. Continuous variables will be checked to determine whether they are normally distributed or not and presented as mean (SD) or median (IQR) and appropriate transformation will be considered for non normally distributed data. Alongside any mixed-model analysis, differences between treatment groups will be presented alongside its 95% confidence interval. An intention-to-treat basis will be used for the primary analysis with per protocol analysis used as part of a follow-up approch. All statistical tests will be 2-sided and significance set at p<0.05.

#### Interim analyses

There will be no formal interim analysis in view of the short recruitment window of the study. Adverse event analysis will be performed after 10 patients have been randomised in each site.

A closed report will be carried out by the study statistician for all independent DMEC meetings. This report will primarily cover quality of data collection and study safety parameters but may also include primary and secondary endpoint data if requested.

### Methods for additional analyses (e.g. subgroup analyses)

#### Primary subgroup analysis

1. Murray Score on randomisation (or pre-ECMO if randomised post-ECMO).
2. COVID-19 vs non COVID-19, based on swab test results (not stratification, binary)
3. ECMO and non-ECMO at randomisation (binary)
4. Comparison of physiological variables in control and intervention groups including: pulmonary shunt, high and low V/Q values, pulmonary mechanics.

#### Exploratory subgroup analysis

5. Focal versus non-focal ARDS (binary)
6. Qualification of driving pressure data as standardised points of clinical management
  1. Driving pressure collected pre-ECMO vs collected post-ECMO
  2. Post-ECMO data only
7. Hyper- and hypo-inflammatory ARDS endotypes

No adjustments are to be made for multiple testing for the above subgroup analysis sets with results considered exploratory in nature with any results interpreted as such.

The Intention-to-Treat (ITT) population will consist of all patients randomised into the DeVENT study. The ITT population will be used for the primary analysis and will be the default population used for any analysis unless otherwise stated.

The Per-Protocol (PP) analysis population set will be based on the ITT population but will exclude all patients under ECMO at randomisation.

In addition to the secondary outcome measures investigating device safety, adverse events and adverse device effects will be summarised by treatment group and severity. A separate table summarising adverse events and their relationship to study treatment will also be produced. In addition, the safety of individual ventilator settings will be assessed. This will include the percentage of time and number of incidences that: tidal volume was higher than 8 ml/kg predicted body weight (PBW) or 10 ml/kg PBW; end tidal carbon dioxide levels were above 7 kPa; SpO_2_ levels were below 90%; for pressure support ventilation, respiratory frequency less than 12 breaths/minute or a rapid shallow breathing index greater than 100 breaths/litre.

### Methods in analysis to handle protocol non-adherence and any statistical methods to handle missing data

Every effort will be made to minimise missing baseline and outcome data in this trial. The level and pattern of the missing data in the baseline variables and outcomes will be established by forming appropriate tables and the likely causes of any missing data will be investigated. This information will be used to determine whether the level and type of missing data has the potential to introduce bias into the analysis results for the proposed statistical methods, or substantially reduce the precision of estimates related to treatment effects. Due to the nature of data being collected, true missing data will only occur in periods where the Beacon device fails to collect data whilst the patient is both non-ECMO and not on control mode ventilation. As the extent of this data-loss is expected to be minimal and is considered missing-at-random we will not carry out any imputation for the primary analysis. A sensitivity analysis will be run to investigate the effect of any missing Beacon data on mean driving pressure values. Data that is not collected due to the patient being on ECMO or support-mode ventilation is not considered to be ‘missing’ and as such would not be considered for any missing-data approaches. No imputation will be considered for the secondary outcome measures

### Plans to give access to the full protocol, participant level-data and statistical code

Details of the trial including study design, eligibility criteria and outcome measures are available to the public on clinicaltrials.gov (NCT04115709). The full SAP is in the online supplement. The trial protocol and SAP are also available on the Imperial College Trials unit website.

### Oversight and monitoring

Imperial Clinical Trials Unit is providing moritoring and oversight for this study for Imperial College London (sponsor). Please see figure 4 for overall reporting structure.

**Figure 4.**
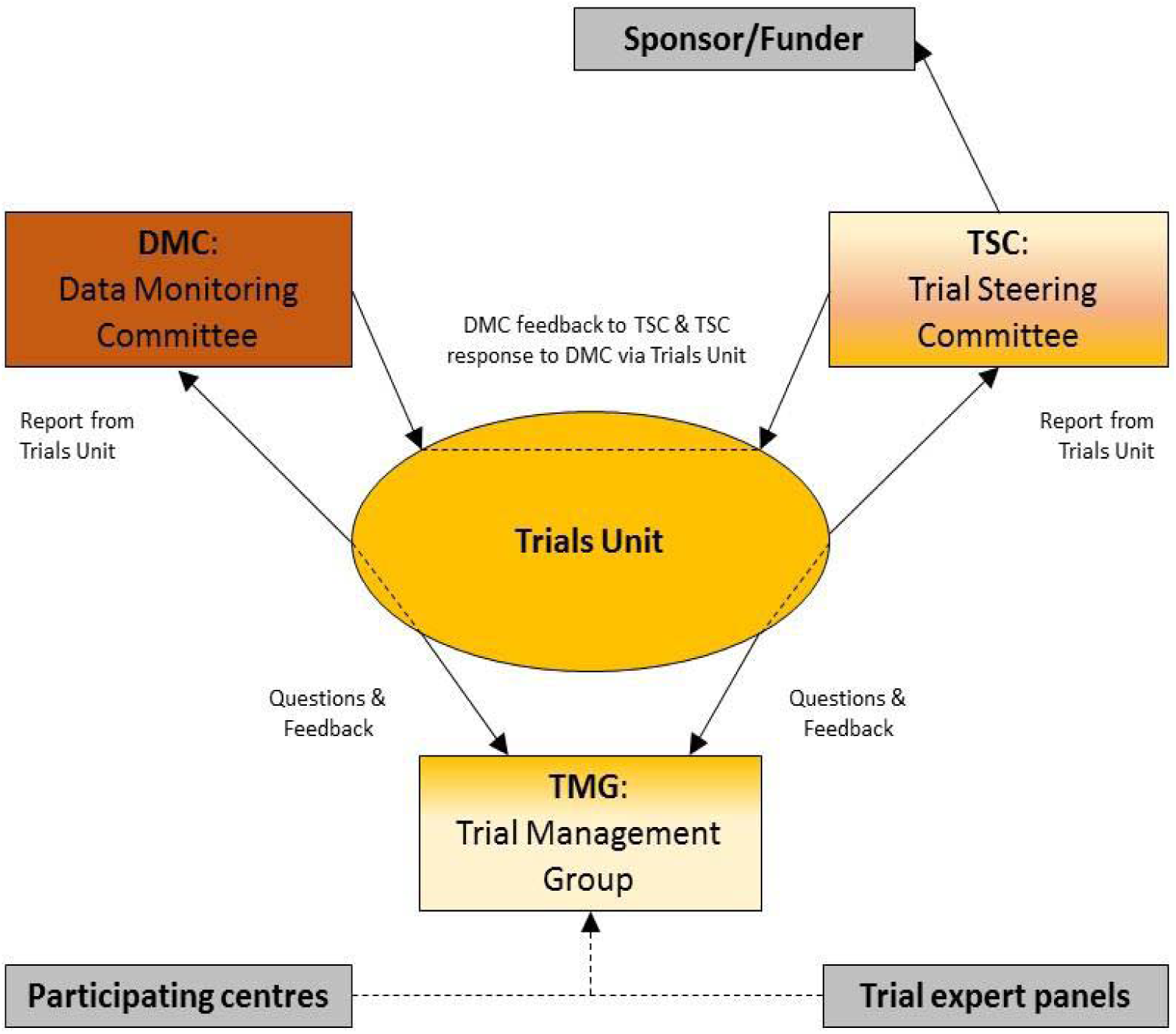
Composition of trial co-ordination

### Composition of the coordinating centre and trial steering committee

Imperial Clinical Trials Unit is the coordinating centre and is a <u>UK Clinical Research Collaboration (UKCRC)</u> registered Clinical Trials Unit. A Trial Steering Committee (TSC) will be convened including as a minimum an independent Chair, independent clinician, the Chief Investigator and Trial Manager. The role of the TSC is to provide overall supervision of trial conduct and progress. Details of membership, responsibilities and frequency of meetings will be defined in a separate Charter.

### Composition of the data monitoring committee, its role and reporting structure

The Data Monitoring and ethics Committee (DMEC) will be comprised of at least 2 independent clinicians, one with experience in clinical trials, and an independent statistician. One of the independent clinicians will have experience in the regulatory aspects of clinical trials involving medical devices. The role of the DMEC will include: monitoring the data and making recommendations to the TSC on whether there are any ethical or safety reasons why the trial should not continue; considering the need for any interim analysis; advising the TSC regarding the release of data and/or information; considering data emerging from other related studies. The DMEC will continually assess both safety and efficacy data on a regular basis with additional meetings convened in the event of any safety concerns. The DMEC should be independent of both the Investigators and the funder/Sponsor and should be the only body that has access to unblinded data. At least two independent members of the DMEC should convene to make the committee quorate.

### Adverse event reporting and harms

It is recognised that the patient population in the ICU will experience a number of common aberrations in physiological values, laboratory values, signs and symptoms due to the severity of their underlying disease and the impact of standard therapies. These will not necessarily constitute an AE unless they require significant intervention, lead to discontinuation of intervention or are considered to be of concern in the investigator’s clinical judgement.

Clinical outcomes from ARDS are exempt from adverse event reporting, <u>unless</u> the investigator deems the event to be related to the use of the device. The following events will be considered clinical outcomes.

- Death related to ARDS and ensuing multi-organ failure
- Neurological insult e.g. intracranial bleeding
- Cardiovascular failure, including the need for vasopressors / inotropes
- Hepatic failure
- Renal failure, including the need for renal replacement therapy
- Haematological / Coagulation failure, including thrombocytopaenia

### Device Deficiency

Definition Device Deficiency:Inadequacy of a medical device related to its identity, quality, durability, reliability, safety or performance, such as malfunction, misuse or use error and inadequate labelling. Device deficiencies include malfunctions, use errors, and inadequate labelling. Any Device deficiencies should be reported to the Device manufacturer (Mermaid Care A/S) as soon as possible after the event, but not longer than 72 hours. Device deficiencies should also be reported in the SMART Trial eCRF, in the form for “Any discontinuation of the Beacon Device”

### Adverse Event (AE) / Adverse Device (ADE) Event

Any untoward medical occurrence in a patient or clinical study subject and which does not necessarily have a causal relationship with this treatment (i.e. any unfavourable or unintended change in the structure (signs), function (symptoms), or chemistry (lab data) in a subject to whom a treatment/study procedure has been administered, including occurrences unrelated to that product/procedure/device).

ADE is an untoward and unintended response to a medical device. The phrase “responses to a medical device” means that a causal relationship between the device under investigation and an AE is at least a reasonable possibility, i.e., the relationship cannot be ruled out.

All cases judged by either the reporting medically qualified professional or the sponsor as having a reasonable suspected causal relationship to the device qualifies as a device effect. This also includes any event resulting from insufficiencies or inadequacies in the instruction for use or deployment of the device and includes any event that is a result of a user error.

### Serious Adverse Events (SAE) / Serious Adverse Device Effects (SADE)

In the event of a Serious Adverse Event (SAE), Serious Adverse Device Effect (SADE) or Unanticipated Serious Adverse Device Effect (USADE) occurring during the subject’s participation in the study the SAE/USADE must be reported to the CI and the Sponsor should be informed. Reporting of the SADE/USADE will also be reported to the Device Manufacturer. For ICTU sponsored studies this will be reported directly to the Joint Research Compliance Office (JRCO) and relevant regulatory authorities where applicable e.g. USADE. A SAE form or eUSADE will need to be completed.

### Serious Adverse Events (SAE)

Any untoward and unexpected medical occurrence or effect that:

- Results in death.
- Is life-threatening – refers to an event in which the subject was at risk of death at the time of the event; it does not refer to an event which hypothetically might have caused death if it were more severe.
- Requires hospitalisation, or prolongation of existing inpatients’ hospitalisation.
- Results in persistent or significant disability or incapacity.
- Is a congenital anomaly or birth defect.
- Is otherwise considered medically significant by the investigator.

Life-threatening” in the definition of “serious” refers to an event in which the subject was at risk of death at the time of the event; it does not refer to an event which hypothetically might have caused death if it were more severe. ** “Hospitalisation” means any unexpected admission to a hospital department. It does not usually apply to scheduled admissions that were planned before study inclusion or visits to casualty (without admission). Medical judgement will be exercised in deciding whether an AE is serious in other situations. Important AEs that are not immediately life-threatening or do not result in death or hospitalisation but may jeopardise the subject or may require intervention to prevent one of the other outcomes listed in the definition above, should also be considered serious.

### Serious Adverse Device Effects (SADE)

A serious adverse device effect (SADE) is any untoward medical occurrence seen in a patient that can be attributed wholly or partly to the device which resulted in any of the characteristics or led to characteristics of a serious adverse event. SADE is also any event that may have led to these consequences if suitable action had not been taken or intervention had not been made or if a circumstance has been less opportune. All cases judged by either the reporting medically qualified professional or the sponsor.

### Unanticipated Serious Adverse Device Effect (USADE)

Any serious adverse device effect on health or safety or any life-threatening problem or death caused by, or associated with a device, if that effect, problem, or death was not previously identified in nature, severity or degree of incidence in the investigational plan or application (including a supplementary plan or application), or any other unanticipated serious problem associated with a device that related to the rights, safety or welfare of the subject.

### Adverse Event recording

AEs will be recorded from the time of consent in the adverse event section of the relevant Case Report Form. All clearly related signs, symptoms, and abnormal diagnostic procedures results should be recorded in the source document using the event terms and grading given in the relevant CRF/eCRF pages. For the purposes of the study, AEs will be followed up according to local practice until the event has stabilised or resolved, or the Follow-up Visit, whichever is the sooner. SAEs will be recorded throughout the study

### Reporting of SAE/SADEs

Reporting of all SAEs (exept common ICU related events) occurring during the study must be performed as detailed in SAE reporting instructions. It is recognised that the patient population in the ICU will experience a number of common aberrations in laboratory values, signs and symptoms due to the severity of their underlying disease and the impact of standard therapies. These will not necessarily constitute an AE unless they require significant intervention, or are considered to be of concern in the investigator’s clinical judgement. For Austrian sites: All serious adverse events must be fully registered by the sponsor and reported immediately to the Federal Office for Health Safety (BASG) and the competent authorities of the other affected contracting parties of the EEA in which the clinical trial is being carried out.

All SAEs will be reviewed by the Chief Investigator or a designated medically qualified representative to confirm expectedness and causality. Reporting of SAEs and review by the CI will be via the trial data collection system (CRF/eCRF).

SAEs that are *related and unexpected*, SADEs and USADEs should be notified to the relevant REC and the Sponsor within 15 days of the Chief Investigator becoming aware of the event. In addition, all SAE/SADE/UADEs should be reported to the manufacturer of the device at the same time. Follow up of patients who have experienced a related and unexpected SAE should continue until recovery is complete or the condition has stabilised. Reports for related and unexpected SAEs should be unblinded prior to submission if required by national requirements.

### Frequency and plans for auditing trial conduct

Quality Control will be performed according to ICTU internal procedures. The study may be audited by a Quality Assurance representative of the Sponsor and/or ICTU. All necessary data and documents will be made available for inspection.

The study may be subject to inspection and audit by regulatory bodies to ensure adherence to GCP and the NHS Research Governance Framework for Health and Social Care (2^nd^ Edition).

### Plans for communicating important protocol amendments to relevant parties (e.g. trial participants, ethical committees)

#### Initial Approval

Prior to enrolment of subjects. The trial will require research and ethical (REC) and Health Research Authority (HRA) approval from all countries. The ethics application made by the Chief Investigator will cover all collaborating sites in the UK. The application to the REC and the relevant NHS R&D offices will be made through the Integrated Research Application System (IRAS). Each EU partner has ethics under its own national / institutional framework.

#### Approval of Amendments

Proposed amendments to the protocol and aforementioned documents must be submitted to the REC and HRA for approval as instructed by the Sponsor. Amendments requiring REC approval may be implemented only after a copy of the REC’s approval letter has been obtained. Amendments that are intended to eliminate an apparent immediate hazard to subjects may be implemented prior to receiving Sponsor or REC approval, refer to urgent safety measures. However, in this case, approval must be obtained as soon as possible after implementation. Annual Progress Reports will be submitted to the Research Ethics Committee (REC) and the Sponsor in accordance with local / national requirements. The Annual Progress Report will also detail all SAEs recorded.

The Sponsor will ensure that the study protocol, Patient Information Sheet (PIS), Informed Consent Form (ICF), GP letter and submitted supporting documents have been approved by the Health Research Authority (HRA) which includes Research Ethics Committee (REC) approval if applicable, prior to any patient recruitment taking place. The protocol and all agreed substantial protocol amendments, will be documented and submitted for HRA approval prior to implementation. All non-UK sites will have documents converted to the host language through official translators.

Before site(s) can enrol patients into the study confirmation of capacity and capability must be issued by the institution hosting the trial. It is the responsibility of the PI at each site to ensure that all subsequent amendments gain the necessary approvals by the participating site. This does not affect the individual clinician’s responsibility to take immediate action if thought necessary to protect the health and interest of individual patients.

### Dissemination plans

We plan to publish our trial protocol and statistical analysis plan to ensure transparency in our methodology. The study findings will be presented at national and international meetings with abstracts on-line. Presentation at these meetings will ensure that results and any implications quickly reach all of the intensive care community. This will be facilitated by our investigator group which includes individuals in executive positions in the UK and European Society of Intensive Care Medicine as well as ECMOnet and EuroELSO committees. In accordance with the open access policies proposed by the NIHR we aim to publish the clinical findings of the trial in high quality peer-reviewed open access (via Pubmed) journals. This will secure a searchable compendium of these publications and make the results readily accessible to the public, health care professionals and scientists.

We will actively promote the findings of the study to journal editors and critical care opinion leaders to ensure the findings are widely disseminated (e.g. through editorials and conference presentations) and are included in future guidelines. Due to limited resources, it will not be possible to provide each patient with a personal copy of the results of the trial. However, upon request, patients involved in the trial will be provided with a lay summary of the principal study findings. The most significant results will be communicated to the public through press releases.

### Trial status

The current protocol attached is version 4, approved on 16^th^ June 2021. The first patient was recruited on 19^th^ March 2020. The last patient was recruited on 4^th^ May 2021 with the data collection continuing with a planned database lock on 30^th^ Sept 2021. Aside from the primary and secondary trial outcomes, the exploratory outcomes will have a final patient visit on 26^th^ March 2022. We were unable to submit this protocol earlier due to clinical pressures as a result of the COVID-19 pandemic.

## Supporting information

Exploratory outcome measures

Final statistical analysis plan

## Data Availability

All data will be available on request through the sponsor.

## Abbreviations

Abbreviation / Acronym: **Full Wording**
ABG: Arterial Blood Gas
AE: Adverse Event
ALPE: Automatic Lung Parameter Estimation
APACHE II: Acute Physiology and Chronic Health Evaluation Score II
APMIC: Anaesthesia, Pain Medicine and Intensive Care
APRV: Airway Pressure Release Ventilation
ARDS: Acute Respiratory Distress Syndrome
BAL: Bronchoalveolar lavage
CI: Chief Investigator
CO_2_: Carbon Dioxide
CONSORT: Consolidated Standards of Reporting Trials
COVID-19: Novel Coronavirus Disease 2019
CRF/eCRF: Case Report Form/Electronic Case Report Form
CTU: Clinical Trials Unit
DMEC: Data Monitoring and Ethics Committee
DNAR: Do Not Attempt Resuscitation
ECMO: Extracorporeal Membrane Oxygenation
ECCO_2_R: ExtraCorporeal Carbon Dioxide Removal
EQ-5D-5L: EuroQoL-5 Dimension Questionnaire (5 level version)
EELV: End-Expiratory lung Volume
FE’CO_2_: End-tidal CO_2_ fraction
FiO_2_: Fraction of Inspired Oxygen
GCP: Good Clinical Practice
HADS: Hospital Anxiety and Depression Score
HRA: Health Research Authority
HRQoL: Health Related Quality of Life
ICF: Informed Consent Form
ICH: International Council on Harmonisation
ICTU: Imperial Clinical Trials Unit
ICU: Intensive Care Unit
IRAS: Integrated Research Application System
MHRA: Medicines and Healthcare products Regulatory Agency
MOCA: The Montreal Cognitive Assessment
NHS: National Health Service
NIHR: National Institute for Health Research
NIV: Non Invasive Ventilation
NMBD: Neuromuscular Blocking Drugs
PaCO_2_: Partial Pressure of Carbon Dioxide in arterial blood
PaO_2_: Partial Pressure of Oxygen in arterial blood
PBW: Predicted Body Weight
PEEP: Positive End Expiratory Pressure
P/F: ratio PaO_2_/FiO_2_ ratio
PI: Principal Investigator
PIS: Patient Information Sheet
PPI: Patient and Public Involvement
Pplat: Plateau Pressure
PTSD: Post Traumatic Stress Disorder
PTSS: 14 Post Traumatic Stress Syndrome Questionnaire
RALE: Radiographic Assessment of Lung Oedema
REC: Research Ethics Committee
RR: Respiratory Rate
SAE: Serious Adverse Event
SAP: Statistical Analysis Plan
SAPS II: Simplified Acute Physiology Score
SBT: Spontaneous Breathing Trial
SGRQ: St George’s Respiratory Questionnaire
SOFA: Sequential Organ Failure Assessment
SOPs: Standard Operating Procedures
SpO_2_: Oxygen Saturation
SRM: Study Reference Manual
TAPSE: Tricuspid Annular Plane Systolic Excursion
TIDIeR: Template for intervention description and replication
TMF: Trial Master File
TMG: Trial Management Group
TSC: Trial Steering Committee
VCO_2_: Carbon Dioxide production
VILI: Ventilator Induced Lung Injury
VO_2_: Oxygen Consumption
Vt: Tidal Volume

## Main protocol contributors

**Dr Brijesh Patel**, Division of Anaesthetics, Pain Medicine, and Intensive Care, Department of Surgery and Cancer, Imperial College, London, UK

**Dr Sharon Mumby**, Airway Disease, National, Heart & Lung Institute, Imperial College, London, UK

**Mr Nicholas Johnson**, Imperial Clinical Trials Unit, Stadium House, 68 Wood Lane, London, W12 7RH

**Dr Emanuela Falaschetti**, Imperial Clinical Trials Unit, Stadium House, 68 Wood Lane, London, W12 7RH

**Mr Jorgen Hansen**, Mermaid Care A/S, Aalborg, Denmark

**Professor Ian Adcock**, Airway Disease, National, Heart & Lung Institute, Imperial College, London, UK.

**Professor Danny F McAuley**, Wellcome-Wolfson Institute for Experimental Medicine, Queen’s University, Belfast, UK

**Professor Masao Takata**, Division of Anaesthetics, Pain Medicine, and Intensive Care, Department of Surgery and Cancer, Imperial College, London, UK

**Dr Dan S**.**Karbing**, Respiratory and Critical Care Group, Department of Health Science and Technology, Aalborg University, Aalborg, Denmark

**Professor Matthieu Jabaudon**, Department of Perioperative Medicine, University Hospital of Clermont-Ferrand, GReD, Université Clermont Auvergne, CNRS, INSERM, Clermont-Ferrand, France

**Associate Professor Peter Schellengowski**, Medical University of Vienna, Department of Medicine I, Waehringer Guertel 18-20, A-1090 Vienna, Austria

**Professor Stephen E**.**Rees**, Respiratory and Critical Care Group, Department of Health Science and Technology, Aalborg University, Aalborg, Denmark

**\*DeVENT study contributors: Imperial College London** – Rhodri Handslip; Sunil Patel; Teresa Lee**; Imperial Clinical Trials Unit** – Ana Boshoff; Mary Cross; **Aalborg University** – Martin S Andersen; **Royal Brompton & Harefield Hospitals** – Mahitha Gummaddi; Vicky Thwaites; Natalie Dormand; Anelise Catelan Zborowski ; Geraldine Sloane; Mathew Varghese; Kamal Liniyage; Adam Pitt; Daniele Cristiano; Rosie Cervera-Jackson; Stephane Ledot; Ben Garfield; James Doyle; Maurizio Passariello; Ben Garfield; Jo Alcada; Alex Rosenberg; **Medical University Vienna** – Thomas Staudinger; Elisabeth Lobmeyr; Nina Buchtele, Oliver Robak, Andja Bojic, Alexander Hermann, Valerie Weinberger, Barbara Krieger, Julia Cserna; **Clermond-Ferrand University Hospital** – Benjamin Rieu, Jean-Baptiste Joffredo, Thomas Costille, Emmanuel Futier, Julien Amat; Dominique Morand; Raiko Blondonnet; **Mermaid Care A/S** – Soren Thomsen; Jakob B Brohus.

**Independent Data Monitoring and Ethics Committee** – Professor Charlotte Summers (Chairperson); Dr Steve Harris; Professor David Harrison (independent statistician); Professor Mark Griffiths

**Independent Trial Steering Committee** – Professor Giacomo Bellani (Chairperson); Professor Dan Brodie; Professor John Laffey; Mrs Yosien Burke (patient and public representative)

## Authors’ contributions

All authors contributed to the development of the protocol. BVP, SM and SR prepared and edited the manuscript. All authors read and approved the final manuscript.

## Availability of data and materials

Not applicable as data is not yet available.

## Ethics approval and consent to participate

The study conduct will comply with all relevant laws of the EU and all relevant laws and statutes of the UK including, but not limited to, the Human Rights Act 1998, the Data Protection Act 1998, the Medicines Act 1968, the Medicines for Human Use (Clinical Trial) Regulations 2004, and with all relevant guidance relating to medicines and clinical studies from time to time in force including, but not limited to, the ICH GCP, the World Medical Association Declaration of Helsinki entitled ‘Ethical Principles for Medical Research Involving Human Patients’ (2008 Version), the NHS Research Governance Framework for Health and Social Care (Version 2, April 2005). In the UK, the study received ethical approval by the London South-East Research Ethics Committee (Ref: 19/LO/1606) on 15 January 2020 with proitocol version 4.0 approved on 16^th^ June 2021. The study protocol was approved by the French Ethics Committee (*Comité de Protection des Personnes Sud Mediterranee III*; approved on July 30, 2020 under number2019.12.06 ter_19.11.15.76132) and Medicine Agency (*Agence Nationale de Sécurité du Médicament*; approved on July 3, 2020 under number 2019-A02610-57-A). The study protocol was approved by the ethics committee of Medical University of Vienna, Austria (EC No. 2056/2019) on 28^th^ April 2020. Consent to enter this study will be obtained after a full account has been provided of its nature, purpose, risks, burdens and potential benefits, with verbal and written explanation and after the patient’s nominated consultee has had the opportunity to consider whether to join the study.

## Consent for publication

Not applicable

