## Supplementary material for "Decision support system to evaluate VENTilation in the Acute Respiratory Distress Syndrome (DeVENT study) – Trial Protocol": Exploratory outcome measures

Supplementary Appendix: Exploratory outcome measures

The pathophysiological and pathobiological processes involved in the progression and resolution of ARDS are poorly understood and there is no current method of predicting trajectory of ARDS. However, the DeVENT study offers a unique opportunity to longitudinally characterise the clinical, physiological, radiological, and biological progression and resolution of ARDS to enable an understanding of disease trajectory. This is currently very important in light of the current COVID-19 pandemic as most ARDS patients is secondary to COVID-19 infection.

**Aims:**

To investigate the clinical, physiological, radiological, and biological mechanisms which characterise the progression and resolution of ARDS (and COVID-19).

**Population:**

All patients recruited to the DeVENT Study in centres that are capable and have the regulatory approvals to attain relevant samples.

**Study assessments:**

Biological Sampling

- **Blood sampling** at (T1) baseline (i.e. on randomisation), and between days 1-3 (T2), 5 -7 (T3), 9-11 (T4), and 13-15 (T5) and weekly (T6, etc) thereafter when in ICU. Blood samples will be taken at subsequent follow-up clinics. Given the urgent nature of COVID-19, a similar sample of blood may be taken prior to starting ECMO (T0) and hence, often prior to discussion with personal nominee, at the discretion of the clinical team. If they are not enrolled into the study then this sample will be discarded.
- **Bronchoscopic sampling** (only if ventilated), with bronchoalveolar lavage (BAL) and deep bronchial brushes, will only be performed if clinically relevant. If patient is in addition on ECMO, patients will be sampled at baseline (T1BAL) and between days 5-7 (T2BAL), and 13-15 (T3BAL) and weekly (T4BAL, etc) thereafter when in ICU. Bronchial brushing maybe performed in some patients. If the patient undergoes a bronchoscopy for clinical purposes, samples will be taken for research purposes dependent on the time of sampling.
- **Urine sampling** (if available) at baseline (T1U) (i.e. on randomisation) and days 1-3 (T2U), 5-7 (T3U), 9-11 (T4U), and weekly (T5U, etc) thereafter.

Excess blood, urine and bronchoscopy samples taken by the clinical team for clinical purposes at the same time-points above may be stored and processed in a similar manner and be utilised for research purposes in order to reduce sampling at any time-points above.

The day of sampling may fall on weekends / holidays when staff are not available. If this does occur, samples/scans will be taken/performed on the day prior/next available day. This will be recorded on the CRF and also on the sample accountability log, along with the day the samples were taken (e.g. day 8 instead of day 7). Blood and urine sampling will be collected by trained study staff and processed according to standardised procedures (1, 2).

Additional blood samples (40ml) may be taken at various time-points for leukocyte isolation. Given the uncertain nature of time of insult, Bronchoscopy and BAL will be undertaken where possible, and processed as previously described (2, 3). Participants will be closely monitored during and after bronchoscopy and BAL. A sample will be sent for microbiological analysis. Some of the following analyses may ALSO be performed in patient and human volunteer samples. From our extensive specialist ICU experience, the expected adverse events associated with bronchoscopy procedures include reversible desaturation and minor contact bleeding.

Biological Analyses

**Transcriptomic, lipidomic, proteomic and metabolomic techniques** may be used on samples such as blood, urine and bronchial brushings to develop clinically useful phenotype handprints. Analyses will be performed to allow intelligent patient and experimental model selection, and identify appropriate samples (for example DNA, RNA, proteins, cells, tissues, blood) for use in the high-dimensional analyses. BAL inflammatory markers (for example cytokines and chemokines measured by ELISA, multiplex sandwich immunoassays, high performance liquid chromatography and meso-scale discovery technology). Breath inflammatory and metabolic markers (including exhaled nitric oxide and metabolomic analysis of volatile organic compounds in exhaled air and metabolites in exhaled breath condensate). Lipid inflammatory mediators may be assessed through the analysis of BAL, serum, plasma and urine biomarkers which may include but not be limited to the measurement of thromboxane B2, prostaglandin E metabolite and 15-epi-lipoxin A4.

To assess **systemic inflammatory and cell death responses**, we may measure plasma and serum inflammatory and cell death response biomarkers which may include but are not limited to measurement of plasma CRP, inflammatory mediators (including but not limited to TNFα, IL1β, IL6, IL8, MLKL, RIPk1/3), proteases and antiproteases, adhesion and activation molecule expression (including but not limited to sICAM1), NETs, coagulation factors (including but not limited to thrombin-antithrombin complex, tissue factor, protein C, thrombomodulin and plasminogen activator inhibitor1), cleaved and whole cytokeratin 16, and Receptor for Advanced Glycation End-products (RAGE) ligands will be undertaken, Specific extracellular vesicle and cellular populations within the blood and BAL (using but not limited to cytospins and flow cytometry) for identification of transcriptome changes within these populations may be carried out.

**Pulmonary inflammatory and cell death responses** will be assessed via BAL biomarkers which may include but are not limited to the measurement of cytokines (including but not limited to TNFα, FasL, IL1β, IL6, IL8), proteases and antiproteases, coagulation factors (including but not limited to thrombin-antithrombin complex, tissue factor, protein C, thrombomodulin and plasminogen activator inhibitor), and RAGE ligands will be undertaken. Identification of specific cellular populations within the BAL (using but not limited to cytospins, flow cytometry, ELISpot assays, in vitro cell expansion) will also be undertaken. Intracellular signalling activity in the alveolar space which may include but not limited to the measurement of BAL total and phosphorylated p38, ERK and JNK MAPKs and STAT -1/-3 from cellular extracts will be measured. Activated and total IκBα and β may be measured in cytoplasmic extracts and NFκβ and AP-1 in nuclear extracts. These investigations will inform further assessment of the influence of clinically relevant causes of weaning failure (e.g. severity of lung pathology, ventilator-induced lung injury and ventilator associated pneumonia).

**Pulmonary and systemic epithelial and endothelial function and injury** may be investigated by testing plasma, serum and BAL biomarkers which may include but are not be limited to measurement of RAGE, cleaved and whole cytokeratin 16, angiopoeitin I/II, surfactant protein-D, von Willebrand Factor, pro-collagen peptide-3 as well as total protein, plasma albumin, α2-macroglobulin, and protein permeability (albumin:α2-macroglobulin ratio) will be undertaken. Urinary albumin/creatinine ratio will also be measured.

Samples from subjects may also be tested on primary cultures of fresh human endothelia, epithelia, neutrophils, monocytes, macrophages and skeletal muscle cells as well as mesenchymal stromal cells to determine surrogate markers of inflammation which may include but not be limited to the measurement of activation (shape change, CD11b surface expression, superoxide release), adhesion and transmigration, cytokine release and matrix metalloproteinase production, rate of apoptosis and their ability to phagocytose. Cells may be isolated from samples for subsequent analysis of cell death and inflammatory pathway activation using a variety of techniques including but not limited to RNA sequencing and flow cytometry.

**Microvesicles** may be isolated from blood and BAL to study the alveolar and systemic microvesicle release, content and function, which may include but not be limited to the measurement of inflammatory mediator release and cell death. Microvesicles may be co-cultured with human cell lines and primary cell cultures in the presence of BAL and serum/plasma from the same patient to determine the effect on their functional properties (cytokine release, phagocytosis, polarization markers expression). Urine micrvovesicles may undergo similar analyses for assessment of renal physiology and dysfunction.

Circulating cells as well as their respective microvesicles may be isolated from blood. Cells may be stimulated (such as monocytes) or matured for 5-7 days to produce monocyte-derived macrophages (MDMs) and stimulated to identify mechanisms which modulate inflammatory responses in these cells. Furthermore, trophic infection may be examined in circulating cells by Flow Assisted Cell Sorting.

**Immunothrombotic analyses** are particularly relevant to COVID-19 given the significant pro-thrombitic nature of this disease. Samples will be utilised for phenotyping ARDS secondary to COVID-19 versus non-COVID-19. Levels of circulating cytokines and vascular dysfunction will be measured as described previously alongside markers of haemostasis (which may include but are not be limited von Willebrand factors (antigen and activity), factor VIII, protein C, protein S, ADAMTS 13, and antithrombin levels) to determine clinical immunothrombotic phenotypes. In addition, we will measure key fibrinolytic parameters involved in the breakdown of clot (which may include but are not be limited fibrinogen and D dimer levels, plasminogen, PAI-1, tPA, soluble thrombomodulin and urokinase levels to determine fibrinolytic capacity and activity). Importantly, ECMO can modulate these coagulation measures and hence, pre-ECMO blood sampling is of paramount importance. Given the low risk of sampling and the high importance to discovering novel therapies to treat COVID-19, this has been added to the sampling regime, and will take place prior to personal nominee advice.

**Physiological analyses**

Daily average physiological status defined as daily averages of measured values of oxygenation (SpO_2_), end-tidal CO_2_ fraction (FE’CO_2_), metabolism (VO_2_, VCO_2_), ventilation (respiratory rate, tidal volume, anatomical dead space), pulmonary mechanics (mean airway pressure, respiratory system compliance), ventilator settings, PaO_2_/FiO_2_, shunt fraction, and end-expiratory lung volume over time as continuously measured by the Beacon system. Proportion of breaths dyssynchronous with the ventilator is to be investgated as an exploratory outcome and compared between groups.

1) An exploratory analysis will be performed to compare parameters describing pulmonary mechanics and gas exchange in COVID and non-COVID patients. This will be performed to investigate whether values of and changes in pulmonary shunt, high V/Q and the mechanical properties of the pulmonary system behave differently in these patient groups. It will be investigating whether the previously postulated pattern of increased shunt and high V/Q without mechanical abnormalities exists within the COVID population.

2) An exploratory analysis will be performed to understand the ventilatory and metabolic measurements performed under ECMO. It will be investigated whether the measurements of oxygen consumption and carbon dioxide production can be used as an estimate of the contribution of the lungs to the exchange of gasses during ECMO, and as such whether an optimal ventilatory strategy can be selected during ECMO so as to minimize lung pressures while preserving pulmonary mechanics and gas exchange.

ICU/hospital cardiac, lung and skeletal/respiratory muscle assessments

a) Echocardiography and lung ultrasound parameters including but not limited to ventricular size and function and tricuspid annular plane systolic excursion (TAPSE), with established clinical protocols.
b) Ultrasound images of the rectus femoris, parasternal muscle and diaphragm will be taken at baseline and on days 1, 3, 7 10, 14 and weekly thereafter, with a final image point on the day of ICU discharge or transfer (or within 24hr of discharge).
c) If able, parasternal and diaphragm imaging will be performed on the first day of a spontaneous ventilation mode.
d) If able, parasternal and diaphragm imaging will be performed during a spontaneous breathing trial (SBT), or when the physician has identified someone as suitable for extubation.
e) If able, parasternal and diaphragm imaging will be performed 24-hours after extubation.
f) Follow-up ultrasound imaging at outpatient follow-up clinics are unchanged and will take place up to 6 and between 6-12 months after ICU discharge.
g) If two time points coincide or are separated by 1 day (i.e. SBT imaging falling on day 7, or first day of spontaneous mode ventilation falling on day 6 etc), then a repeat set of images will not be taken the following day. Rectus femoris imaging time-point will also be adjusted accordingly to avoid repeated disruption to the patient.

ICU/hospital radiological assessments

a) RALE score may be assessed.
b) Axial thoracic CT scans may be assessed for relative proportions of normal lung, ground-glass opacity and lung consolidation and scored as previously described (4) to give a total lung parenchyma score (TLS).
c) Axial thoracic CT scans may be assessed for focal and non-focal patterns of ARDS as previously described (5).
d) Axial thoracic-abdominal CT scans may be assessed for skeletal and respiratory muscle cross-sectional area.

Functional and long-term outcome measures

Variables which impact long-term patient related outcome will be measured in a longitudinal manner:

1. ICU/hospital/follow-up skeletal/respiratory/cardiac muscle ultrasound.
2. ICU/hospital/follow-up imaging (X-ray, CT, ultrasound) and lung function in relation to ARDS (as per usual clinical practice).
3. Sf-36 Health Related Quality of life and EQ-5D-5L (patient and carers) (6 month and 1 year after randomization)
4. St George’s Respiratory Questionnaire (SGRQ) (1 year after randomization)
5. Cognitive Status Questionnaire e.g. mini Mental State examination; PTSS-14; Hospital Anxiety and Depression Scale (HADS) (1 year after randomization)
6. Return to work rates e.g. W&SAS (patient and carers) (6 month and 1 year after randomization)
7. 6-minute walk test (6 month and 1 year after randomization)
