## Supplementary material for "Decision support system to evaluate VENTilation in the Acute Respiratory Distress Syndrome (DeVENT study) – Trial Protocol": Final statistical analysis plan

### Statistical Analysis Plan (SAP)

### **De**cision support system to evaluate **VENT**ilation in **ARDS**

### DeVENT

(IRAS - 266521)

#### Study Investigators:

Chief Investigator: Dr Brijesh Patel

Study Management: Imperial Clinical Trials Unit (ICTU)

#### SAP Working Group:

Emanuela Falaschetti – Senior Statistician

Nicholas Johnson – Study Statistician

| Version | Date | Author |
| --- | --- | --- |
| 1.0 | 13 APRIL 2021 | Nicholas Johnson |

Approved by:

| Name | Signature | Role | Date |
| --- | --- | --- | --- |
| Dr Brijesh Patel | | Chief Investigator | 09 July 2021 |
| Prof. Giacomo Bellani | John Laffey Digitally signed by John Laffey Date: 2021.07.08 20:03:54 +01'00' | TSC Chair | |
| Emanuela Falaschetti | Emanuela Falaschetti Digitally signed by Emanuela Falaschetti Date: 2021.04.16 10:53:02 +01'00' | Senior Statistician | |
| Nicholas Johnson | Nicholas A Johnson Digitally signed by Nicholas A Johnson Date: 2021.07.09 09:46:41 +01'00' | Study Statistician | |

**Decision support system to evaluate VENTilation in ARDS**

**DeVENT**

#### Table of Contents

**2. Study Objectives / Hypotheses Testing** 4
**2.1. Primary Objective** 4
**2.2. Secondary Objective** 4
**2.3. Exploratory Objective** 4
**3. Study Endpoints** 4
**3.1. Primary Endpoint** 4
**3.2. Secondary Endpoints** 4
**3.3. Exploratory Endpoints** 5
**4. DeVENT Study** 5
**4.1. Background/Introduction** 5
**4.2. Study Design** 7
**4.3. Treatment Groups** 7
**4.4. Study Population** 7
4.4.1. Pre-registration evaluations 7
4.4.2. Inclusion Criteria 8
4.4.3. Exclusion Criteria 8
4.4.4. Withdrawal criteria 8
**4.5. Schedule of Time and Events** 9
**4.6. Sample Size Calculation** 13
**4.7. Randomisation** 14
**5. Analysis Sets** 14
**6. Variables of Analysis** 15
**6.1. Baseline Variables** 15
6.1.1. Demographics, medical history and aetiology 15
6.1.2. Day Zero Data 15
6.1.3. Treatment Information 15
**6.2. Primary Endpoint Variable** 15
6.2.1. Associated data for the primary endpoint analysis 16
**6.3. Secondary Endpoint Variables** 16
6.3.1. Beacon Data 16
6.3.2. Clinical Data (via CRF & SMART-TRIAL database) 18
**6.4. Variables for Exploratory Sub-group Analysis** 19
**7. Statistical Methodology** 20
**7.1. Baseline Demographics** 20
**7.2. Primary Analysis** 20
**7.3. Secondary Analysis** 22
**7.4. Per-Protocol Analysis** 23

**7.5. Exploratory Sub-group Analysis** 23
**7.6. Additional Safety Analysis** 23
**8. Statistical Considerations** 24
**8.1. Missing Data and Imputation** 24
8.1.1. Missing Data and Imputation – Primary Outcome Analysis 24
8.1.2. Missing Data and Imputation – Secondary Outcome Analysis 24
**8.2. Adjustments for Multiple Testing** 24
**8.3. Interim Analysis** 24
**8.4. Protocol Deviations** 25
Appendix 1 – Revision History 24
Appendix 2 – References 24
Appendix 3 – Beacon Output for Driving Pressure 27
Appendix 4 – Summary of Output 30
Appendix 5 - Dummy Tables, Figures & Listings 32

#### 2. Study Objectives / Hypotheses Testing

##### 2.1. Primary Objective

To assess the average driving pressure delivered by the mechanical ventilator over the period of time when ARDS ventilation management is advised by the Beacon Caresystem as compared to standard care.

##### 2.2. Secondary Objective

In mechanically ventilated patients with ARDS we will determine the effects of the Beacon Caresystem compared to standard care in a number of outcome measures (see Section 3.2)

##### 2.3. Exploratory Objective

To investigate the clinical, physiological, radiological, and biological mechanisms which characterise the progression and resolution of ARDS (see Section 3.3)

#### 3. Study Endpoints

##### 3.1. Primary Endpoint

The overall final analysis will be performed as the average driving pressure per unit time. However, given the nature of ICU management (transfer, disconnection, nebulisation etc), the driving pressure calculation will only involve periods when the Beacon Caresystem is operating and attached successfully to the patient. Full details of how the driving pressure is collected is detailed in section 6.2 and Appendix 2.

##### 3.2. Secondary Endpoints

**1)** Daily average calculated delivered pressure over time, for periods of spontaneous breathing.
**2)** Daily average calculated mechanical power over time.
**3)** Daily average calculated oxygenation index over time.
**4)** Daily average ventilatory ratio over time.
**5)** Ventilator free days at 90 days.
**6)** Time from control mode to support mode.
**7)** Number of changes in ventilator settings per day.
**8)** % of time in control mode ventilation.
**9)** % of time in support mode ventilation.
**10)** Total duration of mechanical ventilation.
**11)** Tidal volume over time.
**12)** PEEP setting over time.
**13)** Ventilation related complications e.g. pneumothorax and/or pneumomediastinum
**14)** Device malfunction event rate
**15)** Device related adverse event rate
**16)** Number of times the advice from the Beacon system is followed through the duration of the study.

##### 3.3. Exploratory Endpoints

**Exploratory endpoints covering physiological, functional and biological measurements, as well as long-term measurements at 6-months and 1-year, will be covered in a separate analysis plan.**

The Beacon Care system includes a number of physiological models, each of which have been validated separately, and as part for the Beacon Care system. Mathematical models of pulmonary gas exchange have been shown to describe patients during and following surgery (11)(12)(13), in the ICU (14-16), and have been validated against the experimental reference technique for measuring gas exchange (17)(18). Mathematical models of acid-base have been shown to accurately simulate changes in CO2, O2 and strong acid in blood (19), as well as the mixing of blood from different sources (20). Models of respiratory drive have been shown to accurately simulate the effects of changes in support ventilation (21). The Beacon Caresystem has been shown in 4-8 hour periods to reduce levels of inspired oxygen, tidal volume, and pressure support, without detrimental effects (22) and while protecting the respiratory muscles (23).

This means that the advice provided by the system is presented to the clinician. The ventilator settings are then changed by the clinician, and the patient's physiological response to these changes is automatically used by the system to re-tune the models and repeat the process of generating new advice. Patients with severe lung abnormalities such as acute respiratory distress syndrome (ARDS), which often result in small, stiff lungs, are often in control ventilation mode with little or no spontaneous breathing. For these patients, the clinician often increases PEEP to try to recruit units of the lung which are collapsed. This can be difficult, as increasing PEEP may result in elevated lung pressure and hence an increased the risk of lung injury, incomplete expiration and air trapping, and haemodynamic compromise, especially in those with heart failure. The Beacon Caresystem will advise the clinician to optimise PEEP according the measured physiology at that time and hence, will enable the clinician to determine if the patient is a responder or non-responder to the PEEP manoeuvre. PEEP optimisation should enable best ventilation according to personalised press-volume assessments with potential reductions in driving pressure. Importantly, risk is mitigated by the fact that the clinician can always override the advice given.

***Purpose of study***. To compare mechanical ventilation in ARDS patients following advice from the Beacon Caresystem to that of standard routine care to investigate whether use of the system results in a better application of PEEP and driving pressure across all severities and phases of ARDS. In addition, the Beacon Caresystem records breath by breath physiological data including pressure, flow and gas waveforms and hence, we will utilise all raw data collected by the Beacon Caresystem to physiologically characterise the progression and resolution phases of ARDS (figure 1). In parallel to the RCT, biological samples will be taken to enable a combined physiological and biological phenotyping of ARDS. Finally, exploratory measures for long term ICU outcomes and radiological indices will also be examined.

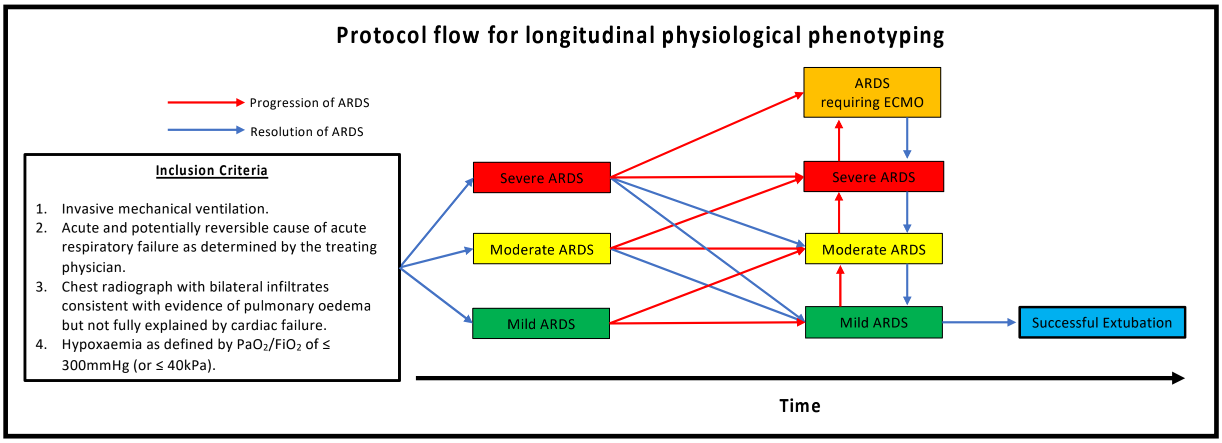

Figure 1: Progression and resolution of ARDS and flow for physiological phenotyping utilising Beacon Caresystem (24).

#### 4.2. Study Design

The DeVENT study is a 2.5-year multi-centre international randomised, controlled, allocation concealed, open, pragmatic clinical trial which will enrol 110 patients with ARDS (according to Berlin definition) with a PaO2/FiO2 of ≤ 300mmHg (or ≤ 40kPa). Patients will be randomised to either have the Beacon Caresystem attached with advice activated (N=55) or standard care with the Beacon Caresystem attached with advice inactive (N=55). The study will also use the Beacon Caresystem to longitudinally physiologically phenotype patients with ARDS.

#### 4.3. Treatment Groups

Patients will be assigned to one of two arms at random:

1. mechanical ventilation in ARDS with advice from the Beacon Caresystem
2. mechanical ventilation in ARDS without advice from the Beacon Caresystem

Full details on the treatment regimen is provided within the study protocol. Further details on the randomisation procedure is provided within Section 4.7

#### 4.4. Study Population

Patients are to be recruited based on the following inclusion/exclusion criteria defined below. Patients are potentially eligible for co-enrolment in other studies, this will be decided on a case-by-case basis in keeping with UK guidelines for critical care research. Imperial Clinical Trials Unit (ICTU) should be informed if co-enrolment is being considered. Co-enrolment with any studies should be documented in the CRF alongside the study identifiers.

##### 4.4.1. Pre-registration evaluations

Patients will only be assessed on the inclusion and exclusion criteria detailed below. This will require a history and examination. An arterial blood gas (ABG) sample and chest x-ray will be needed but would be collected as part of routine clinical care.

##### 4.4.2. Inclusion Criteria

- Invasive mechanical ventilation.
- A known clinical insult with new worsening respiratory symptoms
- Chest radiograph with bilateral infiltrates consistent with evidence of pulmonary oedema but not fully explained by cardiac failure.
- Hypoxaemia as defined by $PaO_2/FiO_2$ of $\leq$ 300mmHg (or $\leq$ 40kPa) (pre-ECMO $PaO_2/FiO_2$ will be used should patient be placed on extracorporeal support).

##### 4.4.3. Exclusion Criteria

- Age < 18 years old.
- The absence of an arterial catheter for blood sampling at study start.
- Consent declined.
- Over 7 days of mechanical ventilation.
- Treatment withdrawal imminent within 24 hours.
- DNAR (Do Not Attempt Resuscitation) order in place
- Severe chronic respiratory disease requiring domiciliary ventilation and/or home oxygen therapy (except for sleep disordered breathing)
- Veno-Arterial ECMO
- Head trauma or other conditions where intra-cranial pressure may be elevated and tight regulation of arterial $CO_2$ level is paramount

##### 4.4.4. Withdrawal criteria

Each participant has the right to withdraw study at any time. In addition, the investigator may discontinue a participant from the study at any time if the investigator considers it necessary for any reason including:

- Ineligibility (either arising during the study or retrospective having been overlooked at screening)
- Significant protocol deviation
- Significant non-compliance with treatment regimen or study requirements
- An adverse event which results in inability to continue to comply with study procedures
- Disease progression which results in inability to continue to comply with study procedures
- Consent withdrawn

To maintain integrity of the randomized trial, all information collected up to that time will still be used and analysed as part of the study.

The reason for withdrawal will be recorded in the eCRF and medical records. Number of withdrawals post-randomisation will be included within the CONSORT diagram.

If the participant is withdrawn due to an adverse event, the investigator will arrange for follow-up visits or telephone calls until the adverse event has resolved or stabilized.

#### 4.5. Schedule of Time and Events

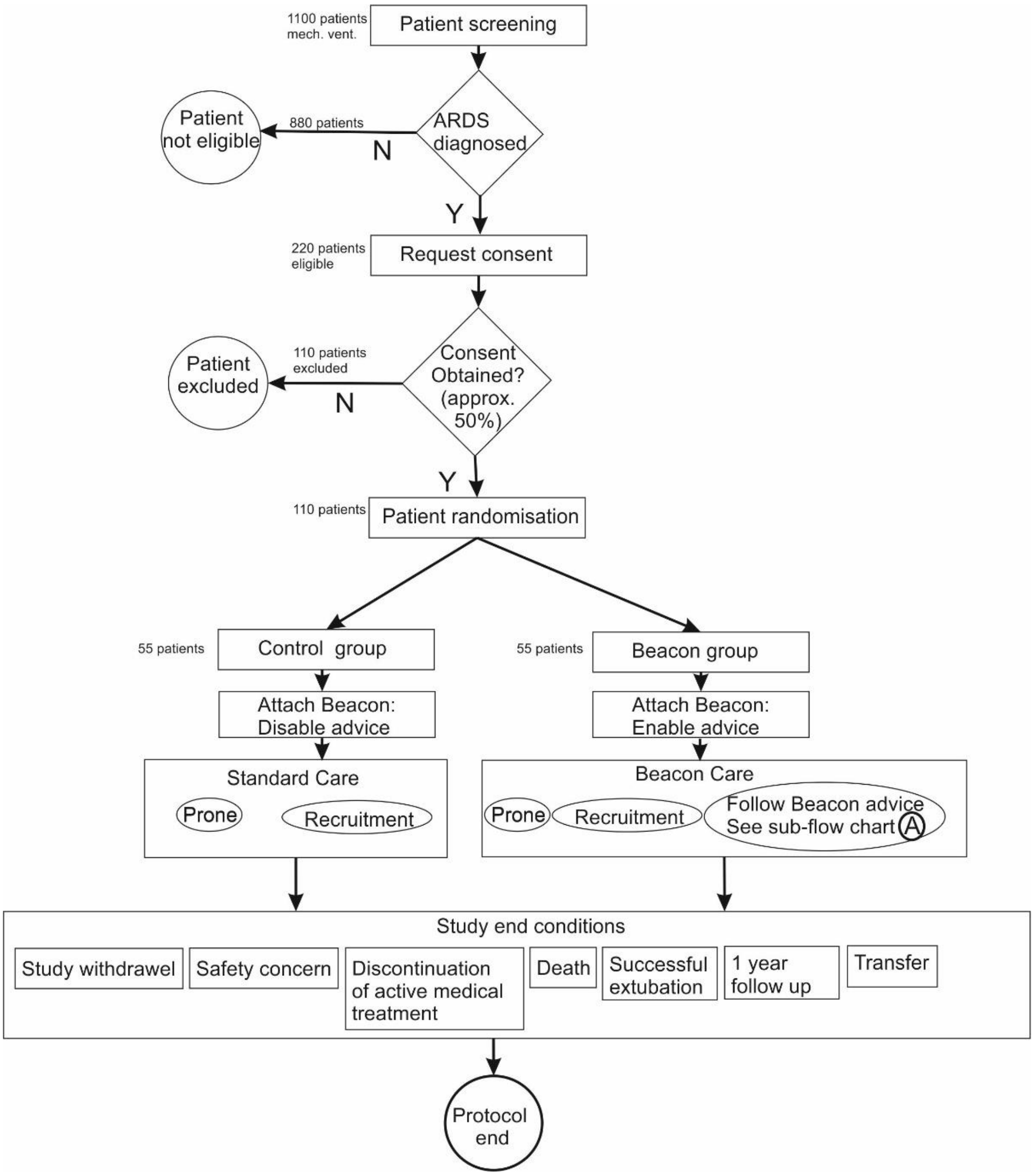

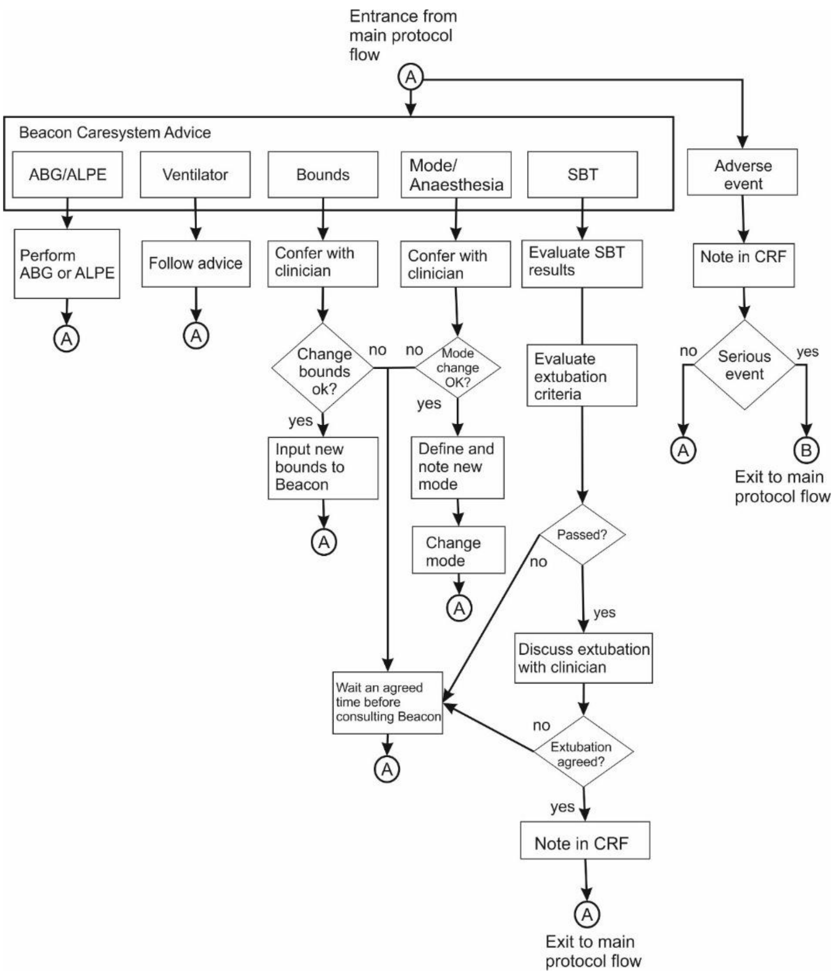

Table 1: Schedule of assessments

| | Days post randomisation | | | | | | | | | | |
|---|---|---|---|---|---|---|---|---|---|---|---|
| | Baseline (0)* | Daily data** | 1 | 2 | 3 | 5 | 7 | 10 | 14 (and weekly thereafter) | Discharge to 6 months (+/- 2 months) | 1 year (+/- 2 months) |
| Consent | X | | | | | | | | | | |
| Inclusion/Exclusion criteria | X | | | | | | | | | | |
| Demographic data | X | | | | | | | | | | |
| Date/time of consent and randomisation | X | | | | | | | | | | |
| Date and time of Hospital and ICU admission | X | | | | | | | | | | |
| Assessment of functional status including co-morbidities (inc. concomitant medication) | X | | | | | | | | | | |
| Smoking history and status | X | | | | | | | | | | |
| Alcohol intake | X | | | | | | | | | | |
| Admission diagnoses | X | | | | | | | | | | |
| Timing of ARDS diagnosis and aetiology | X | | | | | | | | | | |

| | | | | | | | | | |
|---|---|---|---|---|---|---|---|---|---|
| Date/time onset of mechanical ventilation | X | | | | | | | | |
| Use and duration of non-invasive ventilation / nasal high flow prior to intubation | X | | | | | | | | |
| Date and time of worst $PaO_2/FiO_2$ ratio | X | | | | | | | | |
| APACHE II | X | | | | | | | | |
| Determinants of Sequential Organ Failure Assessment (SOFA) score | X | | X | X | X | X | X | X | X |
| Ventilation parameters including but not limited to: Mode of ventilation, minute volume, RR, mean airway pressure, peak inspiratory pressure, plateau pressure, PEEP; I:E ratio; compliance; resistance defined as daily averages of measured values including but not limited to oxygenation ($SpO_2$), end-tidal $CO_2$ fraction ($FE'CO_2$) | X | X | | | | | | | |
| Arterial blood gas including but not limited to $FiO_2$, $PaO_2$, $PaCO_2$, pH | X | X | | | | | | | |
| Prior use of adjunctive therapies including NMBD, prone position, recruitment maneuvers | X | X | | | | | | | |
| Fluid balance – cumulative and daily | X | X | | | | | | | |
| Vasopressor / Inotropic agent usage | X | X | | | | | | | |
| Diuretic usage and use of renal replacement therapy | X | X | | | | | | | |
| Laboratory results in last 24 hours | X | X | | | | | | | |
| Microbiological results to date | X | X | | | | | | | |
| Date/time of attachment of Beacon Caresystem | X | | | | | | | | |
| Commencement or transfer for ECMO following randomization | | X | | | | | | | |
| If patient is on ECMO, then ECMO parameters including but not limited to: ECMO rpm, blood flow, sweep flow, post-oxy, pre-oxy, ECMO 100% test | | X | | | | | | | |
| Daily average physiological status defined as daily averages of measured values including but not limited to oxygenation ($SpO_2$), end-tidal $CO_2$ fraction ($FE'CO_2$), and $PaO_2/FiO_2$ | | X | | | | | | | |
| Daily average physiological status as continuously measured by the Beacon Caresystem | | X | | | | | | | |
| ICU and pre-/post-ICU hospital lung imaging and function in relation to ARDS which includes but not limited to chest X-ray and CT imaging). | | X | | | | | | | |

| | | | | | | | | | | | |
|---|---|---|---|---|---|---|---|---|---|---|---|
| Echocardiography and lung ultrasound parameters including but not limited to ventricular size and function and tricuspid annular plane systolic excursion (TAPSE), with established clinical protocols. | | | X | | X | | X | | X | | |
| Skeletal and respiratory muscle ultrasound (in sites with capability) | | | X | | X | X | X | X | X | X | X |
| Adverse events | | X | | | | | | | | | |
| Is patient fit for extubation and reason for non-extubation (if spontaneous breathing criteria are passed or positive evaluation performed by clinician | | X | | | | | | | | | |
| Sf-36 Health Related Quality of life and EQ-5D-5L (patient and carers) | | | | | | | | | | X | X |
| SGRQ | | | | | | | | | | X | X |
| Cognitive functional tests<br>mini Mental State examination<br>PTSS-14<br>Hospital Anxiety and Depression Scale (HADS) | | | | | | | | | | X | X |
| Return to work rates (W&SAS) (patient and carers) | | | | | | | | | | X | X |
| Primary and Secondary care utilisation | | | | | | | | | | X | |
| 6 minute walk-test | | | | | | | | | | X | X |
| Contact with NHS Digital if required | | | | | | | | | | | X |
| Lung imaging and function in relation to ARDS (as per current clinical protocol i.e. if clinically indicated, which includes pulmonary function tests, CT imaging (including dual energy protocol) | | | | | | | | | | X | X |
| Date and time of any discontinuation of Beacon Caresystem and reason | | | | | | | | | | | X |
| Date and time of any discontinuation of mechanical ventilation | | | | | | | | | | | X |
| Date and time of critical care discharge | | | | | | | | | | | X |
| Date and time of hospital discharge | | | | | | | | | | | X |
| Date and time of death | | | | | | | | | | | X |
| Dates and times of neuromuscular blockade | | | | | | | | | | | X |
| Dates and times of prone position | | | | | | | | | | | X |
| Dates and times of recruitment maneuvers | | | | | | | | | | | X |
| Dates and times of ECMO or $ECCO_2R$ support | | | | | | | | | | | X |

Sample collection:

| | Days post randomisation | | | | | | | | | |
|---|---|---|---|---|---|---|---|---|---|---|
| ***Samples ^*** | Baseline (Day 0)* | 1 | 2 | 3 | 5 | 7 | 10 | 14 (and weekly thereafter) | Discharge to 6 months (+/- 2 months) | 1 year (+/- 2 months) |
| Blood | X | X | X | X | X | X | X | X | X | X |
| Bronchoscopy (only sedated and ventilated) with bronchoalveolar lavage (BAL) | X | | | X | | X | | X | | |
| Bronchoscopy when on ECMO | X | X | X | X | X | X | X | X | | |
| Urine | X | X | | X | | X | | X | X | X |

*24 hours before randomisation. If more than one value is available for this 24-hour period, the value closest but prior to the time of randomisation will be recorded

** Day 1 is from the time of randomisation to the end of that calendar day. If more than one value is available for this period, the value closest but after the time of randomisation will be recorded. All other daily measurements will be recorded and collected between 6-10am or as close to this time as possible, unless otherwise stated in the CRF.

^ The day of sampling may fall on weekends / holidays when staff are not available. If this does occur, samples/scans will be taken/performed on the day prior/next available day. This will be recorded on the CRF and also on the sample accountability log, along with the day the samples were taken (e.g. day 8 instead of day 7)

#### 4.6. Sample Size Calculation

The required sample size for the primary outcome is 110 patients. 55 patients per group will allow to detect a difference of 2 cmH2O in driving pressure between the groups with 90% power and a two-sided alpha of 0.05 assuming a control group driving pressure of 15 cmH2O with a standard deviation of 2.5 cmH2O and including a 40% dropout (33 patients per group for analysis). We have used data from the MIMIC dataset (as published in Serpo Neto et al (25)) for the estimation of the driving pressure. In view of the longitudinal analysis, loss to follow up has taken account of an average mortality of 34% and a 6% drop out.

A follow-up calculation was undertaken in January 2021 to evaluate whether the extent of driving pressure data being collected was sufficient to answer the primary objective. This was due to a higher proportion of patients being exclusively on ECMO than expected due to the COVID-19 outbreak.

Basing the calculation on a repeated-measures basis using estimates of the intraclass correlation co-efficient (ICC) of driving pressure values and the estimate of the coefficient of variation (CV) for days data collected (per patient) we calculated the expected patients required to fulfil the primary outcome. Using the data extract collected for the second DMEC meeting (December 2020, n=38) we estimated values for ICC and CV of 0.3 and 0.8 respectively. Based on these estimates and by providing alternative scenarios we were able to create the following table:

| | | ICC | | |
|---|---|---|---|---|
| CV | | **0.1** | **0.3** | **0.5** |
| | **1.0** | 12 | 19 | 23 |
| | **0.8** | 11 | 17 | 22 |
| | **0.6** | 11 | 16 | 21 |

From the above, under the 'worst-case' scenario; with a CV of 1.0 and an ICC of 0.5 we would require 23 patients per arm to provide sufficient driving pressure data to have a powered analysis. Based on the December 2020 data it is estimated that approximately 96 patients would be required for a powered analysis. A further investigation (February 2021, n=65) confirmed that the estimates for ICC and CV were appropriate and were unlikely to go beyond the extreme values presented above.

Comparing this to the original calculation and taking the increased rate of ECMO patients into consideration, it is expected that the original proposal to randomise 55 patients per group would allow us to collect sufficient driving pressure data to answer the primary objective.

##### 4.7. Randomisation

Patients will be randomly allocated to one of two groups, i.e. the Standard Care group or the Beacon group. A separate randomisation list will be created for each site and stratified by ECMO/non ECMO and COVID/non-COVID. Patients will be centrally allocated to an arm of the study from a master randomisation list (one per site) created by an independent statistician. Given the unblinded nature of this device study, allocation concealment will be maintained through random size block allocation of patients (however due to the potential low numbers in some strata, block sizes are restricted to ensure balancing).

Full details can be found in the Study Specific Procedure Manual (SSPM) for Randomisation (Version 1.1, 31st May 2020)

#### 5. Analysis Sets

The Intention-to-Treat (ITT) population will consist of all patients randomised into the DeVENT study. The ITT population will be used for the primary analysis and will be the default population used for any analysis unless otherwise stated.

The Per-Protocol (PP) analysis population set will be based on the ITT population but will exclude all patients under ECMO at randomisation.

### 6. Variables of Analysis

#### 6.1. Baseline Variables

##### 6.1.1. Demographics, medical history and aetiology

The following data will be collected at enrolment and stored in the SMART-TRIAL database management system.

1. Patient demographics – Age, Gender, Ethnicity, Height, Weight, Predicted Weight, Ulna Length
2. Smoking and Alcohol History
3. Comorbidities – CPD, CVD, Metabolic & Endocrine, Neurological, and Immunosuppression
4. Use and duration of non-invasive ventilation / nasal high flow prior to intubation
5. Days since ARDS onset
6. ARDS aetiology
7. Medications – Steroids (acute & chronic), statins, PPIs, ace inhibitors, etc…
8. Patient Vasopressor / Inotrope agent usage
9. Patient Diuretic usage and use of renal replacement therapy
10. Acute Physiology and Chronic Health Evaluation score (APACHE II)
11. Determinants of Sequential Organ Failure Assessment (SOFA) score
12. Microbiological results to date

##### 6.1.2. Day Zero Data

13. Ventilation parameters including but not limited to: Mode of ventilation, minute volume, Respiratory Rate (RR), mean airway pressure, peak inspiratory pressure, plateau pressure, PEEP; I:E ratio; compliance; resistance and measured values including but not limited to oxygenation ($SpO_2$), end-tidal $CO_2$ fraction ($FE'CO_2$).
14. Arterial blood gas including but not limited to $FiO_2$, $PaO_2$, Partial Pressure of Carbon Dioxide in arterial blood ($PaCO_2$), pH
15. Prior use of adjunctive therapies including Neuromuscular Blocking Drugs (NMBD), prone position, recruitment manoeuvres
16. Fluid balance – cumulative and daily up to day 0
17. Date and time of worst $PaO_2/FiO_2$ ratio
18. Laboratory results (in last 24 hours).

##### 6.1.3. Treatment Information

The following information will be presented in a listing as supplementary information

19. Date/time of consent and randomisation
20. Date and time of hospital and ICU admission
21. Date/time of onset of mechanical ventilation
22. Date/time of attachment of Beacon Caresystem

#### 6.2. Primary Endpoint Variable

**Measured Driving Pressure (via inspiratory/expiratory pauses)**

Via Beacon Device

The primary endpoint variable of driving pressure will be collected via the Beacon device and will be derived using the Beacon data and stored externally. Further information detailing Beacon output can be found in Appendix 1. Driving pressure will be measured during inspiratory pauses greater than 1 second and will be derived using measured respiratory pressures at the end of inspiratory (Pplat) and expiratory (PEEP) pauses:

$$DP = Pplat - PEEP.$$

Due to the extent of mechanical assistance provided under periods of ECMO and on ventilator support mode, evaluable driving pressure values can only be collected whilst the patient is a) not on ECMO and b) ventilator setting is on control mode.

In the event where more than one driving pressure value has been measured within a study day, the mean will be calculated and used as the driving pressure value for that particular day.

Collection at Site (Recorded in CRF)

Driving pressure will also be measured daily (where possible) by the clinical team with end inspiratory and expiratory pause times recorded in the CRF alongside measured Pplat and PEEP. An intrinsic PEEP value will also be derived based on the measured PEEP minus the set PEEP on the ventilator. These values will be used to validate the values collected at a similar time by the Beacon device. In the event where no corresponding Beacon value has been collected within a study day (due to device deficiency events, as defined in section 8.5) the value recorded in the CRF will also be used in the primary analysis.

Further information detailing Beacon output can be found in Appendix 2.

##### 6.2.1. Associated data for the primary endpoint analysis

Alongside driving pressure, the following variables will be used in the mixed-model defined in section 7.2:

- Treatment Group (assigned at randomisation)
- ECMO status at randomisation
- COVID status at randomisation
- Study site (where patient is) enrolled
- Total number study days where patient is non-ECMO and not on support mode
- Duration of ventilation prior to connection (1 to 7 days).
- Duration of hospital admission prior to intubation

#### 6.3. Secondary Endpoint Variables

##### 6.3.1. Beacon Data

1. Daily average calculated delivered pressure over time, for periods of spontaneous breathing in non-ECMO periods.

   Alongside paused inspiratory/expiratory driving pressure data, the Beacon device will also record, on a continuous breath-by-breath basis, a surrogate driving pressure derived using peak pressure and set PEEP. The corresponding surrogate driving pressure for each study day will then be derived using the daily mean.

2. Daily average calculated mechanical power over time in non-EMCO periods.

   Mechanical Power is derived from the Beacon data on a minute-by-minute basis. Any day where the patient is in non-EMCO and within control mode, a daily mean for mechanical power will be produced for analysis.

3. Daily average calculated oxygenation index over time.

   Oxygenation Index is a composite endpoint derived from the following:

   - $F_iO_2$ = fraction of inhaled oxygen (%)
   - $P_{AW}$ = mean airway pressure (mmHg)
   - $PaO_2$ = Partial pressure of arterial oxygen (mmHg)

   Any day where the patient is in non-EMCO, the daily mean Oxygenation Index will be calculated using the derived values for oxygenation index, where Oxygenation Index equals:

$$\frac{F_iO_2 \text{ x } P_{AW}}{P_aO_2}$$

4. Daily average ventilatory ratio over time

   Ventilatory ratio is a measure of impaired ventilation. Ventilatory ratio is defined as:

$$\frac{\text{minute ventilation (ml/min)} \times P_aCO_2\text{(mmHg)}}{\text{predicted body weight (kg)} \times 100 \times 37.5}$$

   Any day where the patient is in non-EMCO, a daily mean for Ventilatory ratio will be produced for analysis.

5. Time from control mode to support mode in non-EMCO periods.

   All switches between control and support modes are to be recorded by the Beacon device. This is defined as where a mode has changed for more than 30 minutes.

6. Number of changes in ventilator settings per day in non-EMCO periods.

   As recorded by the Beacon device

7. % of time in control mode ventilation in non-EMCO periods

   Device mode will be monitored throughout by the Beacon device. Percentage of overall time in control mode will be reported under non-ECMO periods only.

8. % of time in support mode ventilation in non-EMCO periods.

   Device mode will be monitored throughout by the Beacon device. Percentage of overall time in control mode will be reported under non-ECMO periods only.

9. Tidal volume over time.

   Tidal volume to be recorded on a minute-by-minute basis of which the mean (SD) tidal volume per study day is to be derived and used for analysis

10. PEEP setting over time.

As recorded by the Beacon device, mean PEEP per study day is to be derived and used for analysis

11. Device malfunction event rate

Beacon device deficiencies will be recorded via the periods where the device is not able to data. Reason for disconnection will be recorded in the SMART-TRIAL database. Rate will be established as minutes per day disconnected whilst subject is connected to the device.

12. Number of times the advice from the Beacon system is followed through the study.

As recorded by the Beacon device. A daily rate will be established based on number of events recorded over time (in days) where the device is connected.

###### 6.3.2. Clinical Data (via CRF & SMART-TRIAL database)

13. Total duration of mechanical ventilation.

Start time and stop time of mechanical ventilation to be recorded and stored in the trial database.

14. Ventilator free days at 90 days.

Ventilator-free days at 90-days post-randomisation will be established using the dates of ventilation collected above and the recorded date of extubation.

15. Ventilation related complications e.g. pneumothorax and/or pneumomediastinum

Based on adverse events recorded and stored within the trial database

16. Device related adverse event rate & serious adverse event rate

Based on adverse events recorded and stored within the trial database

#### 6.4. Variables for Exploratory Sub-group Analysis

*Primary subgroup analysis:*

1. Murray Score on randomisation (or pre-ECMO if randomised post-ECMO).
2. COVID-19 vs non COVID-19, based on swab test results (not stratification, binary)
3. ECMO and non-ECMO at randomisation (binary)
4. Comparison of physiological variables in control and intervention groups including: pulmonary shunt, high and low V/Q values, pulmonary mechanics.

*Exploratory subgroup analysis:*

5. Focal versus non-focal ARDS (binary)
6. Qualification of driving pressure data as standardised points of clinical management
    a. Driving pressure collected pre-ECMO vs collected post-ECMO
    b. Post-ECMO data only
7. Hyper- and hypo-inflammatory ARDS endotypes

#### 7. Statistical Methodology

##### 7.1. Baseline Demographics

Patient characteristics will be presented via a table of summary statistics. Summaries of continuous variables will be presented as means and standard deviations if normally distributed, and as medians and inter-quartile ranges for skewed data. Categorical variables will be presented as frequencies and percentages.

##### 7.2. Primary Analysis

All randomised patients to be considered for the primary analysis but will need to have at least one recorded driving pressure value (measured from inspiratory/expiratory pauses) to be included.

The primary outcome will be analysed using a mixed-model approach. Due to the varying extent of time periods where patients are non-EMCO and not on pressure support some patients will have more days with evaluable driving pressure than others. A random effect for patient intercept will also be included in the regression model and will take the following form:

$$Y_{ij} = b_{0\mathrm{i}} + \beta_1 X_{1i} + \mathrm{xxxxx} + \beta_\mathrm{n} X_{\mathrm{n}i} + e_{\mathrm{ij}}$$

$Y_{ij}$ refers to the score on the dependent variable for patient *i* collected at study day *j*

$b_{0i}$ refers to the random intercept per patient *i where* $b_i \sim \mathrm{N}(0,D)$

$\beta_1 X_1$ refers to the fixed effect (slope) for variable $X_1$

$\beta_\mathrm{n} X_\mathrm{n}$ refers to the fixed effect (slope) for variable $X_n$ where *n* represents number of variables in the model

$e_{ij}$ refers to the random error for patient *i* at visit *j* where $e_{\mathrm{ij}} \sim \mathrm{N}(0,\sigma^2)$

**Let $DP_{ij}$ represent the mean driving pressure for patient *i* from readings collected at study day *j*.**

We will model then $DP_{ij}$ as the sum of 9 components:

$$DP_{ij} = \mathrm{intercept}_i + \beta_1.\mathrm{treatment}_i + \beta_2.\mathrm{NENS\_Days}_i + \beta_3.\mathrm{ECMO}_i + \beta_4.\mathrm{COVID}_i + \beta_5.\mathrm{Site}_i + \beta_6.\mathrm{clinical1}_i + \beta_7.\mathrm{clinical2}_i + \mathrm{residual\ error}_{ij}$$

1. intercept term: represents the random intercept term for patient *i* and is assumed to have a normal distribution with mean 0 and standard deviation *D*
2. treatment term: the primary outcome, represents treatment group assigned to patient i where 0 = no Beacon and 1 = Beacon
3. NENS_Days: number of study days where patient is non-ECMO and not on support mode for patient *i*

4. ECMO term: ECMO status for patient i at randomisation where 0 = no ECMO and 1 = ECMO
5. COVID term: COVID status for patient i at randomisation where 0 = no COVID and 1 = COVID
6. Site term: Site at which patient i is enrolled where 0 = RBH, 1 = MUV, 2 = UCA
7. Clinical Term 1: Duration of ventilation prior to connection (1 to 7 days).
8. Clinical Term 2: Duration of hospital admission prior to intubation
9. residual error term is assumed to have a normal distribution with mean 0 and standard deviation σ2

$\beta_1$ to $\beta_8$ represents the fixed effects coefficients for the 8 independent variables defined above.

The above model will be run to the following specifications:

- Fixed Effects: Treatment, ECMO-status, COVID-status (all binary), Site (categorical; 3-levels), total days non-ECMO,non-support (discrete), clinical 1 & clinical 2 (as above)
- Random Effects: Intercept (per patient)
- Covariance Structure: Unstructured*
- Estimation Method: ML
- Computation Method: Type III Tests of Fixed Effects

* If model fails to converge an alternative structure may be used.

Analysis will be carried out using PROC MIXED in SAS or using the corresponding procedural command in an alternative software package. The code will take the following form:

```
PROC MIXED DATA=FinalAnalysisData METHOD=ML;
  CLASS treatment ECMO COVID site;
  MODEL DP = treatment time_vent ECMO COVID Site Clin1 Clin2 /s CL
RESIDUAL ;
  RANDOM intercept;
run;
```

The model assumptions, including normality, will be evaluated using residual and other diagnostic plots of model fit and sensitivity analyses using alternative methods (e.g., transformation) will be considered if there appear to be substantial deviations from model assumptions.

**The primary outcome investigating the effect of Beacon device on the mean driving pressure will be tested based on the estimate of the fixed effect for treatment group (defined as $\beta_1$ above). The estimated treatment effect will be reported alongside its 95% confidence interval. All other model effects will also be reported. Statistical tests will be two-tailed with a 5% significance level.**

Mean driving pressure values will also be presented in a summary table showing means and standard deviations if normally distributed, or as medians and inter-quartile ranges if skewed. If transformed, the transformed data will also be presented in a summary table alongside the mixed-model output. Data will be grouped by treatment and stratification variables; ECMO status, COVID-19 status and site.

#### 7.3. Secondary Analysis

The following daily average values and continuous outcome variables will be assessed using the same mixed model as defined in section 7.2 above

- Daily average calculated delivered pressure over time, for periods of spontaneous breathing.
- Daily average calculated mechanical power over time.
- Daily average calculated oxygenation index over time.
- Daily average ventilatory ratio over time
- Total duration of mechanical ventilation.
- % of time in control mode ventilation
- % of time in support mode ventilation.
- Device related adverse event rate

The following daily average values and continuous outcome variables will be assessed using the mixed model as defined in section 7.2 but with additional fixed parameters added for study_day (of measurement) and the interaction term between treatment*study_day. A random effect for study day will also be incorporated.

- Tidal volume over time.
- PEEP setting over time.

The remaining secondary endpoint variables will be presented via summary statistics per treatment group for continuous data or by frequency table if categorical (details below).

- Time from control mode to support mode.

  Median (IQR) time in days to (first) change from control to support mode will be presented for each treatment group alongside the difference between groups with 95% CI

- Ventilator free days at 90 days.

  Median (IQR) ventilator-free days will be presented for each treatment group alongside the difference between groups with 95% CI

- Ventilation related complications e.g. pneumothorax and/or pneumomediastinum

  Frequency of events for each treatment group will be presented by event type and by overall events. Comparison of overall between the two groups done using the chi-squared or Fishers exact test (where total events in one or both arms $\leq 5$)

- Number of times the advice from the Beacon system is followed.

  Frequency of events for each treatment group will be presented

- Number of changes in ventilator settings per day (under support mode).

Frequency of events for each treatment group will be presented.

- Device malfunction event rate

  Frequency of events for each treatment group will be presented overall and broken down by type of device deficiency. Median (IQR) time lost to device deficiency will be presented by group.

##### 7.4. Per-Protocol Analysis

The primary analysis will be re-run using the per protocol population defined in Section 5 (only non-ECMO patients). The same methodology will be used as per section 7.2 but an additional covariate for baseline driving pressure will be added.

##### 7.5. Exploratory Sub-group Analysis

*Primary subgroup analysis:*

The primary analysis will be re-run with an additional sub-group covariate and their interaction with treatment group added for the following scenarios:

1. Murray Score on randomisation (or pre-ECMO if randomised post-ECMO).
2. COVID-19 vs non COVID-19, based on swab test results (not stratification, binary)
3. ECMO and non-ECMO at randomisation (binary)
4. Physiological variables as below:
    a. Pulmonary shunt (continuous)
    b. High vs low V/Q values (binary)
    c. Pulmonary mechanics

*Exploratory subgroup analysis:*

Data permitting, the primary analysis may be re-run with an additional sub-group covariate and their interaction with treatment group added for the following scenarios:

1. Focal versus non-focal ARDS (binary)
2. Qualification of driving pressure data as standardised points of clinical management
    a. Driving pressure collected pre-ECMO vs collected post-ECMO (binary)
    b. Post-ECMO data only
3. Hyper- and hypo-inflammatory ARDS endotypes (binary)

##### 7.6. Additional Safety Analysis

In addition to the secondary outcome measures investigating device safety, adverse events and adverse device effects will be summarised by treatment group and severity. A separate table summarising adverse events and their relationship to study treatment will also be produced.

In addition, the safety of individual ventilator settings will be assessed. This will include the percentage of time and number of incidences that: tidal volume was higher than 8 ml/kg predicted body weight (PBW) or 10 ml/kg PBW; end tidal carbon dioxide levels were above 7 kPa; $SpO_2$ levels were below 90%; for pressure support ventilation, respiratory frequency less than 12 breaths/minute or a rapid shallow breathing index greater than 100 breaths/litre.

#### 8. Statistical Considerations

##### 8.1. Missing Data and Imputation

Every effort will be made to minimise missing baseline and outcome data in this trial. The level and pattern of the missing data in the baseline variables and outcomes will be established by forming appropriate tables and the likely causes of any missing data will be investigated. This information will be used to determine whether the level and type of missing data has the potential to introduce bias into the analysis results for the proposed statistical methods, or substantially reduce the precision of estimates related to treatment effects. If necessary, these issues will be dealt with using multiple imputation based on the rules below:

###### 8.1.1. Missing Data and Imputation – Primary Outcome Analysis

Due to the nature of data being collected, true missing data will only occur in periods where the Beacon device fails to collect data whilst the patient is both non-ECMO and not on control mode ventilation. As the extent of this data-loss is expected to be minimal and is considered missing-at-random we will not carry out any imputation for the primary analysis. A sensitivity analysis will be run to investigate the effect of any missing Beacon data on mean driving pressure values.

Data that is not collected due to the patient being on ECMO or support-mode ventilation is not considered to be ‘missing’ and as such would not be considered for any missing-data approaches.

###### 8.1.2. Missing Data and Imputation – Secondary Outcome Analysis

No imputation will be considered for the secondary outcome measures presented in section 6.3

##### 8.2. Adjustments for Multiple Testing

No adjustment for multiple testing will be made for secondary endpoints using the same mixed-model method as section 7.2.

All sub-group analysis incorporating the mixed-model method of section 7.2 will be considered exploratory in nature and must be interpreted as such.

##### 8.3. Interim Analysis

Interim reports detailing baseline demographics, safety variables and the primary endpoint (if requested) will be produced for assessment and discussion at DMEC meetings, the framework of which will be provided within the DMEC charter.

Other than the above, no formal interim analyses are planned. Additional reports may be requested by the DMEC or TSC if necessary.

#### 8.4. Protocol Deviations

Protocol deviations are to be listed and summarised by category and site.

### APPENDICIES

#### Appendix 1: Revision History

| Version Number | Date | Comments |
|---|---|---|
| 0.0 | 11 March 2021 | Final draft for TSC/DMEC review |
| 0.1 | 24 March 2021 | Revisions following TSC Review (additional info to sample size, additional safety analysis) |

#### Appendix 2: References

8. Zampieri FG, Costa EL, Iwashyna TJ, Carvalho CRR, Damiani LP, Taniguchi LU, et al. Heterogeneous effects of alveolar recruitment in acute respiratory distress syndrome: a

machine learning reanalysis of the Alveolar Recruitment for Acute Respiratory Distress Syndrome Trial. British journal of anaesthesia. 2019 Apr 5.

#### Appendix 3: Beacon Output for Driving Pressure

Each patient will have a report provided by the Beacon team which will consist of a plot similar to the one shown in figure App 1 below:

Figure App 1: Example patient Beacon Report

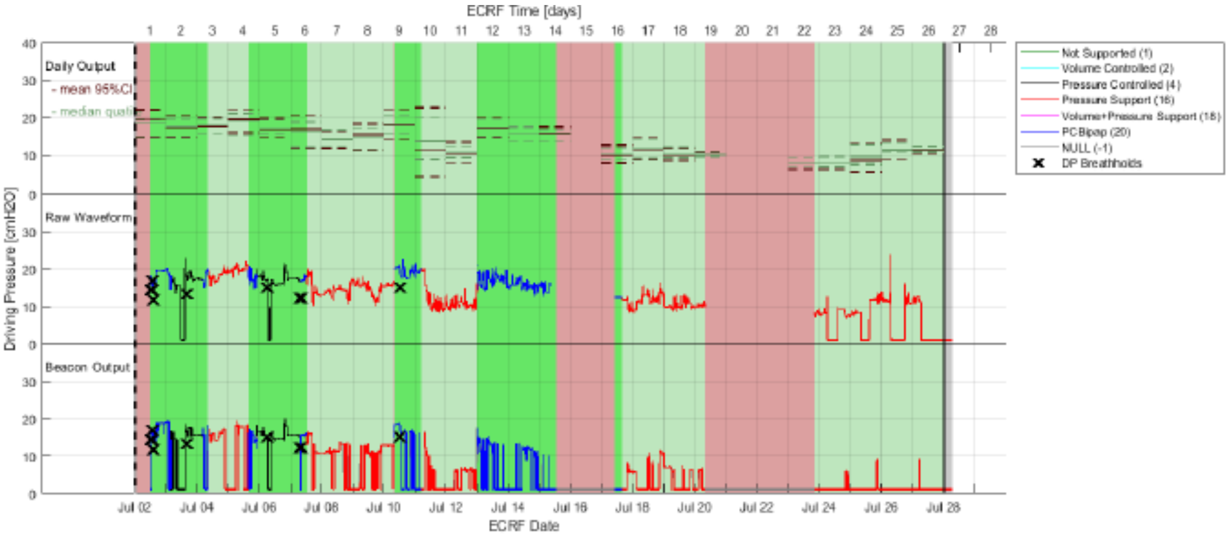

The key sections of this report are as follows:

The background colour scheme represents the periods of data collection. Dark green indicates when the patient is non-EMCO and also not on pressure support. It is in these dark green sections where assessable primary endpoint data can be collected.

The black crosses represent driving pressure breathholds. In other words, where the driving pressure has been measured via an inspiratory/expiratory pause. Any black cross contained in a dark green section will represent primary endpoint data.

Alongside the black crosses there are three sections representing 'waveform' pressure data. The lower 'Beacon' output plot represents data derived via the Beacon device. The middle plot represents the 'raw' waveform data. The colour of both lines represents the ventilator mode at that particular time with red indicating the patient is on pressure support and blue where the patient is on control mode.

The top plot shows graphically the summary stats of the waveform data for each study day. The corresponding mean values will be offset to derive the surrogate driving pressure values as per section 6.2.2.

#### Derivation of driving pressure:

To calculate the driving pressure the following needs to be performed on the pressure wave forms:

1) Identify breaths with an inspiratory pause – these are those with a very long inspiratory time (defined as greater than 1 second)
2) Read the end inspiratory pressure at the end of the pause
3) Read the end expiratory pressure at the end of the subsequent breath.
4) Subtract these two pressures

The Beacon team have developed code to automatically go through each breath data and calculate the driving pressure.  An example can be seen for a single breath below.

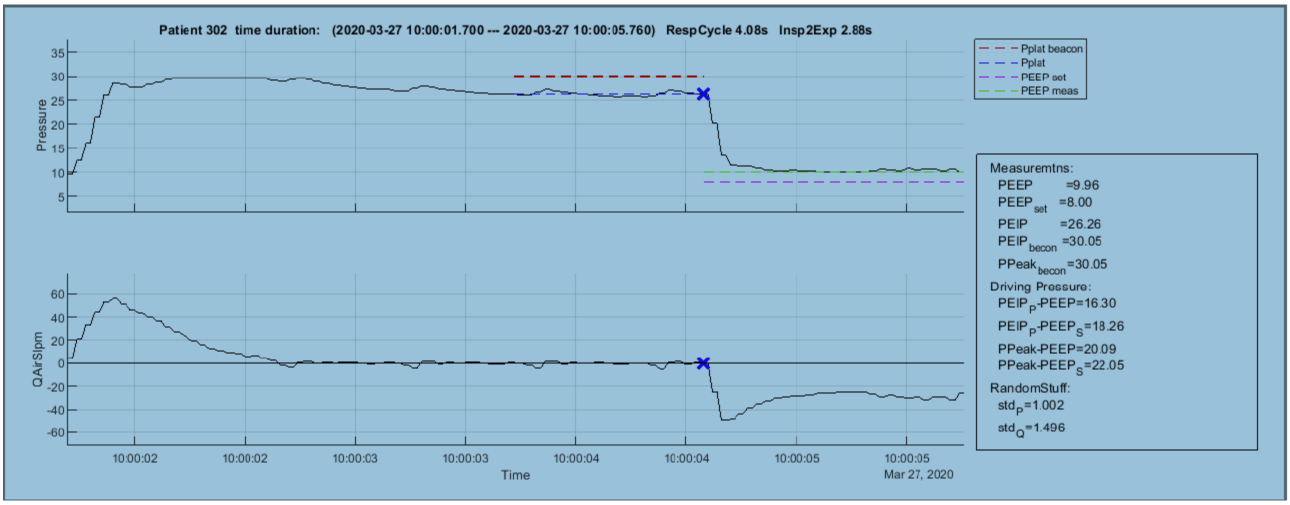

The top curve is pressure and the bottom flow. The long inspiration is identified and the blue cross is the end inspiratory pause pressure. The green line is the end expiratory pressure.

For this analysis we can provide you complete documents of all paused breaths per patient (typically two or more per day) plus a table of the driving pressure at time points for each patient. You should note that if the patient is on ECMO or in a pressure support mode the above calculations are not possible/relevant.

In addition to the above, it is possible to get a surrogate driving pressure on every breath, by assuming the peak inspiratory pressure is almost equivalent to the pause. This is represented by the following plot

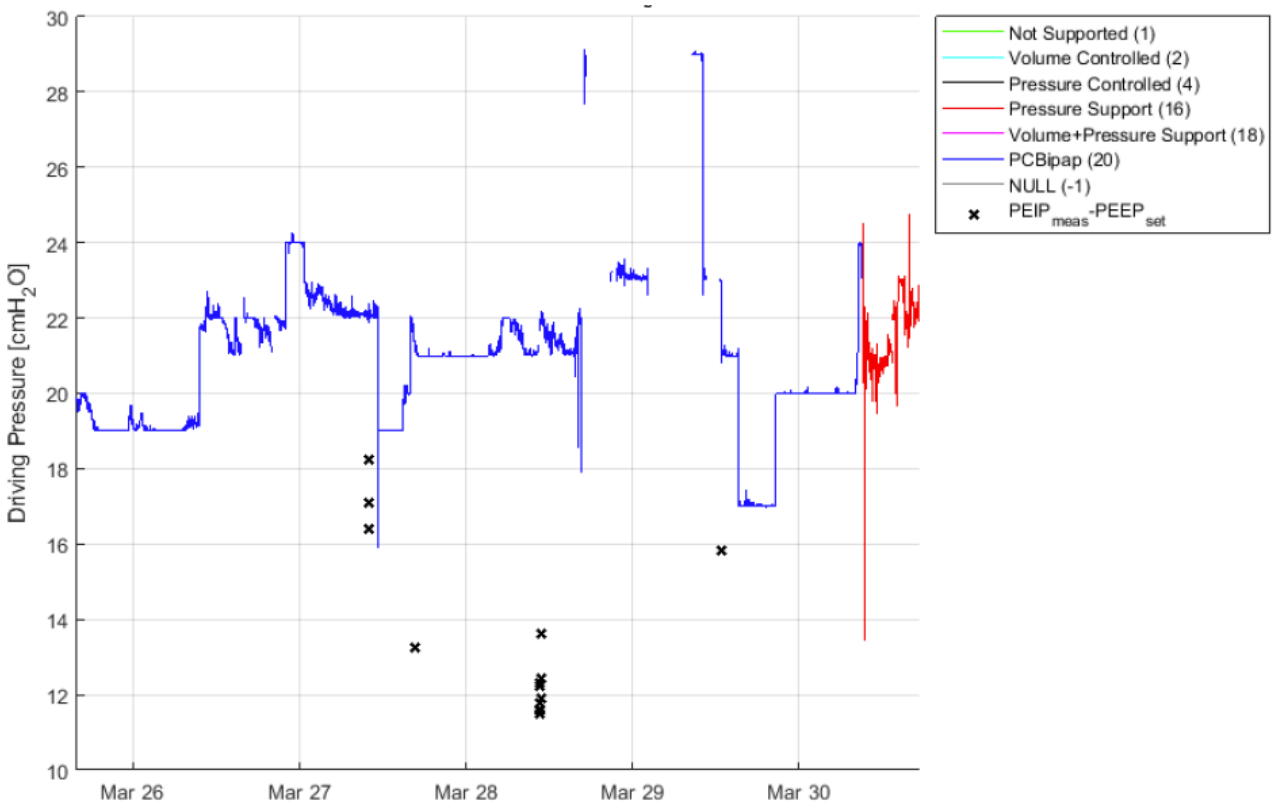

The blue curve will be the surrogate driving pressure over all days, with the black crosses the driving pressure on individual days. The red is a period of pressure support which cannot be used.

#### Appendix 4: Summary of Output

| DeVENT Output Index | | |
|---|---|---|
| Ref No | Title | Template |
| 0.1 | Summary of Randomisation by Site | 1 |
| 0.2 | Summary of Randomisation by Strata (COVID & ECMO) | 2 |
| 0.3 | Flowchart of Data (CONSORT Diagram) | 3 |
| 0.4 | Summary of Clinical (non-device) Related Deficiencies resulting in loss-of-data | 4 |
| 0.5 | Summary of end-of-protocol events | 5 |
| 1.1 | Baseline Demographics | 6 |
| 1.2 | Day Zero Data | 6 |
| 2.1 | Summary of Driving Pressure Data | 7 |
| 2.2 | Primary Analysis of Driving Pressure Data | 8 |
| 2.3 | Summary of Driving Pressure Data - Per Protocol | 7 |
| 2.4 | Analysis of Driving Pressure Data - Per Protocol | 8 |
| 3.1.1 | Summary of daily average calculated delivered pressure over time | 7 |
| 3.1.2 | Analysis of daily average calculated delivered pressure over time | 8 |
| 3.2.1 | Summary of daily average calculated mechanical power over time. | 7 |
| 3.2.2 | Analysis of daily average calculated mechanical power over time. | 8 |
| 3.3.1 | Summary of daily average calculated oxygenation index over time. | 7 |
| 3.3.2 | Analysis of daily average calculated oxygenation index over time. | 8 |
| 3.4.1 | Summary of daily average ventilatory ratio over time | 7 |
| 3.4.2 | Analysis of daily average ventilatory ratio over time | 8 |
| 3.5.1 | Summary of Total duration of mechanical ventilation. | 7 |
| 3.5.2 | Analysis of Total duration of mechanical ventilation. | 8 |
| 3.6.1 | Summary of Percentage of time in control mode ventilation | 7 |
| 3.6.2 | Analysis of Percentage of time in control mode ventilation | 8 |
| 3.7.1 | Summary of Percentage of time in support mode ventilation | 7 |
| 3.7.2 | Analysis of Percentage of time in support mode ventilation | 8 |
| 3.8.1 | Summary of Proportion of breaths dysyncronous with the ventilator. | 7 |
| 3.8.2 | Analysis of Proportion of breaths dysyncronous with the ventilator. | 8 |
| 3.9 | Summary of Device malfunction event rate | 7 |
| 3.10.1 | Summary of Device related adverse event rate | 7 |
| 3.10.2 | Analysis of Device related adverse event rate | 8 |
| 3.11.1 | Summary of changes in tidal volume over time. | 7 |
| 3.11.2 | Analysis of changes in PEEP setting over time. | 8 |
| 3.12.1 | Summary of changes in PEEP setting over time. | 7 |
| 3.12.2 | Analysis of changes in PEEP setting over time. | 8 |
| 3.13 | Summary of Time from control mode to support mode | 7 |
| 3.14 | Ventilator free days at 90 days | 7 |
| 3.15 | Ventilation related complications e.g. pneumothorax and/or pneumomediastinum | 7 |
| 3.16 | Number of times the advice from the Beacon system is followed. | 7 |
| 3.17 | Number of changes in ventilator settings per day (under support mode). | 7 |
| 4.1 | Subgroup analysis of Murray Score on randomisation | 8 |
| 4.2 | Subgroup analysis of COVID vs non-COVID (as per swab testing) | 8 |
| 4.3 | Subgroup analysis of ECMO and non-ECMO at randomisation | 8 |

| DeVENT Output Index | | |
|---|---|---|
| Ref No | Title | Template |
| 4.4.1 | Subgroup analysis of Pulmonary Shunt Fraction | 8 |
| 4.4.2 | Subgroup analysis of High vs low V/Q values | 8 |
| 4.4.3 | Subgroup analysis of Pulmonary mechanics | 8 |
| 4.5 | Subgroup analysis of Focal versus non-focal ARDS | 8 |
| 4.6.1 | Subgroup analysis of Driving pressure collected pre-ECMO vs collected post-ECMO | 8 |
| 4.6.2 | Analysis of Driving Pressure for post-ECMO data | 8 |
| 4.7 | Subgroup analysis of Hyper- and hypo-inflammatory ARDS endotypes | 8 |
| 5.1 | Summary of Adverse events by Severity | 9 |
| 5.2 | Summary of Adverse events by Relationship to Study | 10 |
| 5.3 | Summary of Device Related Adverse events by Severity | 9 |
| 5.4 | Summary of Adverse events by Relationship to Device | 10 |
| 5.5 | Summary of Serious Adverse events by Severity | 9 |
| 5.6 | Summary of Serious Adverse events by Relationship to Study | 10 |
| 5.7 | Summary of Serious Device Related Adverse events by Severity | 9 |
| 5.8 | Summary of Serious Adverse events by Relationship to Device | 10 |
| 5.9 | Incidences of Tidal Volume events | 11/12 |
| 5.10 | Incidences of High end-tidal Carbon Dioxide events | 11/12 |
| 5.11 | Incidences of Low $SpO_2$ levels | 11/12 |
| 5.12 | Incidences of respiratory/breathing events | 11/12 |
| 6.1 | Listing of Protocol Deviations | 13 |
| 6.2 | Listing of Protocol Violations | 13 |

### Appendix 5: Dummy Tables, Figures & Listings

##### Template 1: Summary of Randomisation by Site

| Site | Beacon | No Beacon | Total |
|---|---|---|---|
| RBH | | | |
| MUV | | | |
| UCA | | | |
| All Sites | | | |

RBH = Royal Brompton Hospital

MUV = Medical University Vienna

UCA = CHU-Clermont Ferrand

##### Template 2: Summary of Randomisation by Strata (COVID & ECMO)

| | | ECMO | | No-ECMO | | |
|---|---|---|---|---|---|---|
| **COVID** | **Site** | **Beacon** | **No Beacon** | **Beacon** | **No Beacon** | **TOTAL** |
| No COVID | All Sites | | | | | |
| COVID | All Sites | | | | | |
| TOTAL | All Sites | | | | | |

**Template 3: Flowchart of Data (CONSORT Diagram)**

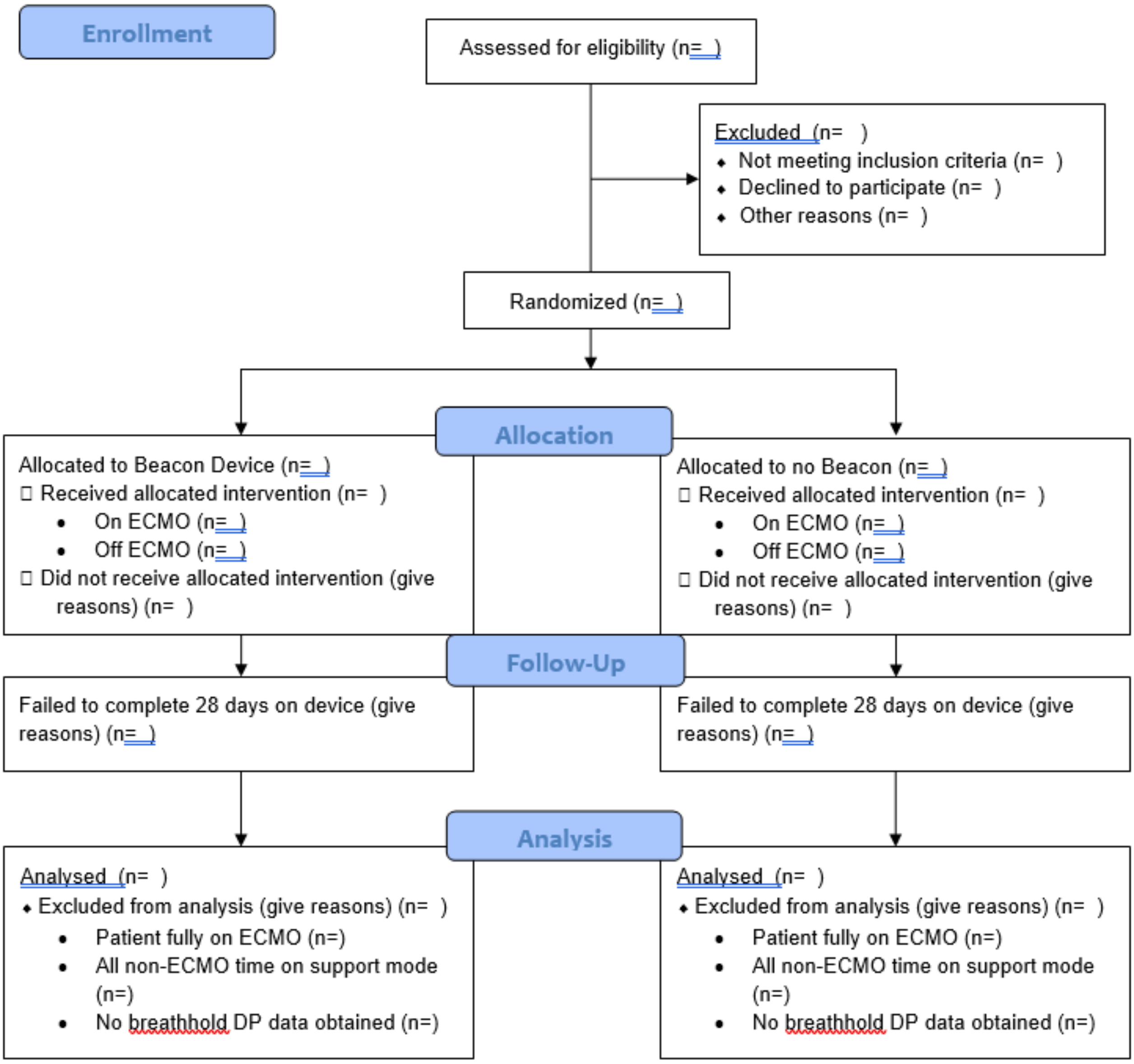

**Template 4: Summary of Clinical (non-device) Related Deficiencies resulting in loss-of-data**

| Category | ECMO | Non-ECMO |
|---|---|---|
| Change of Ventilator<br>…<br>Other (Loss of Power) | | |

* Above events are only for periods where the subject is not on ECMO.

**Template 5: Summary of end-of-protocol events**

| Category | Number of occurrences | Median [IQR] study days to event |
|---|---|---|
| Extubation<br>Transfer to another hospital<br>Patient Death<br>… | | |

**Template 6: Baseline Demographics**

| Variable | Statistics | ECMO | No-ECMO | All |
|---|---|---|---|---|
| Age at Consent (y) | Mean (SD) | | | |
| Ethnicity (%) | Asian | | | |
| | Black | | | |
| | Hispanic | | | |
| | Indian Subcontinent | | | |
| | White | | | |
| Gender – Male (%) | n (%) | | | |
| Height (cm) | Mean (SD) | | | |
| Weight (kg) | Mean (SD) | | | |
| BMI (kg/m2) | Mean (SD) | | | |
| Smoking History (%) | Current smoker | | | |
| | Unknown | | | |
| | Never Smoker | | | |
| | Ex-Smoker | | | |
| Comorbidities – CPD (%) | n (%) | | | |
| | Unknown | | | |
| Comorbidities – CVD (%) | n (%) | | | |
| | Unknown | | | |
| Comorbidities – Metabolic & Endocrine | n (%) | | | |
| | Unknown | | | |
| Comorbidities – Chronic Immuno-suppression | n (%) | | | |
| | Unknown | | | |
| Comorbidities – Chronic neurological disease | n (%) | | | |
| | Unknown | | | |
| Days since Ards Onset | Median [IQR] | | | |

**Template 7: Summary Statistics**

| Variable* | Treatment | n | Mean | SD | Median | IQR | Min. | Max. |
|---|---|---|---|---|---|---|---|---|
| xxxxx | Beacon | | | | | | | |
| | No Beacon | | | | | | | |

**Template 8: Mixed-Model Output**

| Effect | Value | Estimate | Standard Error | DF | t Value | Pr > \|t\| | Alpha | Lower | Upper |
|---|---|---|---|---|---|---|---|---|---|
| **Intercept** | | | | | | | | | |
| **treatment** | Beacon | | | | | | | | |
| **treatment** | No Beacon | | | | | | | | |
| **ECMO** | ECMO | | | | | | | | |
| **ECMO** | No-ECMO | | | | | | | | |
| **VISITORDER** | | | | | | | | | |
| **…** | | | | | | | | | |
| **…** | | | | | | | | | |

| Type 3 Tests of Fixed Effects | | | | |
|---|---|---|---|---|
| **Effect** | **Num DF** | **Den DF** | **F Value** | **Pr > F** |
| **treatment** | | | | |
| **ECMO** | | | | |
| **…** | | | | |
| **…** | | | | |

**Template 9: AE Frequency Table for Severity**

| | | Beacon | No Beacon | Total |
|---|---|---|---|---|
| Severity | Mild | | | |
| | Moderate | | | |
| | Severe | | | |
| | Total | | | |

**Template 10: AE Frequency Table for Relationship**

| | Subjects with AEs* | | | | | | |
|---|---|---|---|---|---|---|---|
| Treatment | No Relationship | Unlikely | Possible | Probable | Definitely | Undefined | Total |
| Beacon | | | | | | | |
| No Beacon | | | | | | | |
| All subjects | | | | | | | |

**Template 11: Listing of Events**

| Treatment | Subject | Site | Type | Start Date | End Date | Value Recorded |
|---|---|---|---|---|---|---|

**Template 12: Frequency of Events**

| | | Beacon | No Beacon | Total |
|---|---|---|---|---|
| Event | Xxx | Xx (xx%) | | |
| | xxx | | | |
| | xxx | | | |
| | Total | | | |

**Template 13: Listing of Protocol Deviations / Violations**

| Treatment | Subject | Site | Type | Start Date | End Date |
|---|---|---|---|---|---|
